# Comparison of approaches to control for intracranial volume in research on the association of brain volumes with cognitive outcomes

**DOI:** 10.1101/2023.07.14.23292678

**Authors:** Jingxuan Wang, Tanisha Hill-Jarrett, Peter Buto, Annie Pederson, Kendra D. Sims, Scott C. Zimmerman, Michelle A DeVost, Erin Ferguson, Benjamin Lacar, Yulin Yang, Minhyuk Choi, Michelle R. Caunca, Renaud La Joie, Ruijia Chen, M. Maria Glymour, Sarah F. Ackley

## Abstract

**Background:** Consistent methods for evaluating the link between brain structure and cognition are essential for understanding determinants of neurologic outcomes. Studies examining associations between brain volumetric measures and cognition use various statistical approaches to account for variation in intracranial volume (ICV). It is unclear if commonly used approaches yield consistent results.

**Methods:** Using a brain-wide association approach in the MRI substudy of UK Biobank (N=41,964; mean age=64.5 years), we used regression models to estimate the associations of 58 regional brain volumetric measures with eight cognitive outcomes, comparing no correction and five ICV correction approaches. Approaches evaluated included: no correction; dividing regional volumes by ICV, with and without further adjustment for ICV (proportional approach); including ICV as a covariate in the regression (adjustment approach); and regressing the regional volumes against ICV in different normative samples and using calculated residuals to determine associations (residual approach). We used Spearman-rank correlations and two consistency measures to quantify the extent to which associations were inconsistent across ICV correction approaches for each possible brain region and cognitive outcome pair across 2,784 regression models.

**Findings:** The adjustment and residual approaches typically produced similar estimates, which were inconsistent with results from the crude and proportional approaches. Inconsistencies across approaches were largest when estimates from the adjustment and residual approaches were further from the null. That is, the approach used was least important when the association between brain volume and cognitive performance was close to null; in this case, all approaches tend to estimate a null association.

**Interpretation:** Commonly used methods to correct for ICV yield inconsistent results and the proportional method diverges from other methods. Adjustment methods are the simplest to implement while producing biologically plausible associations.

## Introduction

Structural magnetic resonance imaging (MRI) is widely used to evaluate the relationship between brain volumetric measures and cognitive measures, including memory, attention, and executive function.^1,2^ In studies examining associations between brain volumetric measures and cognition in aging populations, correcting volumetric measures for intracranial volume (ICV) is often necessary to account for differences in skull size.^3,4^ This is because skull size is not an independent predictor of dementia^5^ and is associated with numerous childhood and adulthood socioeconomic status factors that may confound associatons between neuroimaging measures and outcomes.^6^ Several different statistical approaches to account for ICV have been adopted in the field. These different approaches may produce inconsistent estimates, but, to date, there is no clear guidance on which approaches are preferable. This presents significant challenges for reproducibility in neuroimaging studies, may account for inconsistent results across studies, and likely contributes to incorrect estimates in some studies. Associations between volumetric measures and treatment or outcomes are used in a wide range of contexts, including drug trials,^7^ and may be used inform biological understanding of disease, research priorities, and interventions.^8^ Thus, analysis decisions on ICV correction may ultimately impact individual clinical diagnosis and management.^9^

The proportional, adjustment, and residual approaches are the three commonly used approaches to correct for ICV (Box 1). For the proportional approach, each brain volume measure is divided by ICV and this scaled quantity is used to determine associations with a cognitive measure, typically using a regression model.^4,10,11^ For the adjustment approach, ICV is included along with the brain volume measure as an independent variable in a regression model with cognition as the dependent variable.^4,11^ The residual approach uses two regression models, first regressing each regional volume against ICV, calculating the residuals from this model (i.e., variation in brain volume not predicted by ICV), and using the calculated residuals as the independent variable in a regression with cognitive measures as the predictor. Typically, but not always, the coefficients for the first regression are estimated in a normative sample of healthy controls.^4,11,12^ Specific implementations of these methods vary, and sometimes crude volumes are used without correcting for ICV.^13^

### Box 1

Summary of commonly used approaches used to correct for intracranial volume (ICV).

| Approach | Description |
| --- | --- |
| *Crude, No Correction for ICV* | Crude volumes are used in determining associations with cognitive outcomes. ICV is not controlled for in either calculating correlations or performing regressions. |
| *Proportional* | Brain volume measures are divided by ICV, and this scaled quantity is used to determine associations with a cognitive measure, typically using a regression model. |
| *Adjustment* | ICV is included along with brain volume measures as an independent variable in a regression model with cognition as the dependent variable. |
| *Residual* | The residual approach uses two regression models, first regressing regional volumes against ICV, calculating the residuals from this model, and using the calculated residuals as the independent variable in a regression with cognitive measures as the predictor. Typically, coefficients for the first regression are estimated in a normative sample of healthy controls. |

Evidence on how different correction approaches modify the estimated associations between MRI volumetric measures and cognition is limited. Some prior studies examine how other associations with MRI volumetric measures, such as sex, gender, and age,^14–16^ vary with ICV correction strategy. The small number of prior studies evaluating how different ICV correction strategies affect associations between volumetric measures and cognition have important limitations: they do not evaluate all commonly used ICV correction approaches; they use a single or small number of cognitive measures; are performed in younger samples or small samples; or they are performed in highly select volunteer cohorts (e.g. the Alzheimer’s Disease Neuroimaging Intitiative cohort) such that results may not generalize to the general aging population.^4,14–16^

In this study, we compared estimated associations between MRI volumetric measures and cognitive measures across commonly used ICV correction approaches in the MRI subsample of the UK Biobank. The UK Biobank is a cohort of middle-aged and older adults participating in the National Health Service. As such, Alzheimer’s disease and vascular dementia would be expected to be the most common causes of brain atrophy.^17,18^ Inspired by brain-wide association studies (BWAS), which evaluate each pairwise association of brain region and outcome,^19^ we evaluated the extent to which different ICV correction approaches give inconsistent associations between a brain volume and cognitive measure for the full-factorial combination of ICV correction approach, brain volumetric measure, and cognitive outcome. Inconsistency across ICV correction approaches may have important implications for reproducibility in neuroimaging research. These inconsistencies may account for conflicting findings across studies and produce spurious associations that would not reach statistical significance with alternative approaches.

## Methods

### Study Population

The UK Biobank is a large prospective cohort study of 502,490 UK adults aged 40-69 years at recruitment in 2006-2010. At the baseline visit, participants completed social, physical, and medical assessments. Starting in 2014, participants were invited for MRI neuroimaging at four clinics using identical protocols.^20^ The final target sample for the MRI substudy is 100,000 but at the time of writing, MRI imaging data were available for 41,964 participants. Ethical approval was obtained by the UKB study from the National Health Service National Research Ethics Service with all participants providing written informed consent. Analyses were approved by the University of California, San Francisco Institutional Review Board under UK Biobank Resource project #74748.

### MRI Volumetric Measures

All MRI imaging was carried out using Siemens Skyra 3 Tesla scanner with a standard 32-channel head coil. Full details on the image acquisition, processing, and quality control are available at UK Biobank Brain Imaging Documentation.^21^ All image preprocessing was conducted by the UK Biobank neuroimaging team and included non-brain removal, bias-field correction, and tissue segmentation. The T1-weighted images with a cubic millimeter isotropic resolution were previously analyzed with FMRIB Software Library (FSL). T1-weighted images and T2-weighted fluid attenuation inversion recovery (FLAIR) images were further processed using FreeSurfer to generate subcortical volumes using the automatic subcortical segmentation (ASEG), and cortical volumes based on the Desikan-Killianny-Tourville (DKT) atlas. We included 27 subcortical and 31 cortical regions (**Supplementary Material 1**). ICV was estimated from ASEG. In our primary analysis, we *a priori* selected eight regions of interest (ROIs) previously linked to cognitive outcomes.^22–26^ Only data from the first MRI visit was used due to limited follow-up. We combined hemispheres to obtain a single measure for each ROI.

### Cognitive Measures

We considered eight cognitive measures: fluid intelligence, numeric memory, prospective memory, pairs matching, Trail Making A, Trail Making B, reaction time, and symbol digit substitution. Detailed descriptions of the eight cognitive tests are included in **Supplementary Material 2.** All cognitive tests were administered in English via touch screen interface and designed to be completed without supervision. All cognitive outcomes were measured at the MRI visit. Cognitive scores for pairs matching, Trail Making A, Trail Making B, and reaction time referred to negative one times the score on those cognitive tests; that is, these measures were signed such that higher values indicate better cognition. All continuous cognitive outcomes were *z*-standardized.

### ICV Correction Approaches

We considered no correction and five implementations of widely used approaches to correct for ICV in analyses of associations between regional brain volumes and cognition (in all models, the cognitive measure is the outcome variable of interest).^10,11,27^ The first approach uses crude, uncorrected volumes, while the other five are ICV correction approaches. The approaches are as follows: 1) **Crude approach:** Crude volumes are used without correcting for ICV to determine associations with cognitive measures in a regression model. 2) **Proportional approach:** volumetric measures are divided by ICV and this ratio of regional volume to ICV is used to determine associations in a regression model with cognition as the outcome. 3) **Proportional with adjustment approach:** The regional volume to ICV ratio from the proportional approach is used to determine associations in a regression model that additionally adjusts for ICV as a covariate. 4) **Adjustment approach:** Crude volumes are used to determine associations in a regression model that adjusts for ICV as a covariate. 5) **Full-sample residual approach:** Each volumetric measure is first regressed against ICV in the full sample of participants. This regression model is then used to obtain ICV-corrected volumes in the whole sample using the following: 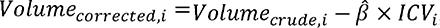, where 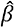 is the slope from the regression model. ICV-corrected volumes (i.e., residuals from the first regression model) are then used as the independent variable in a regression model to determine associations with cognitive measures. 6) **Normative-subsample residual approach**: This approach is identical to the full-sample residual approach, but the first regression is restricted to a “normative” sample of dementia-free participants younger than 60 years.

### Covariates

Covariates included age, sex^14,15^ (female, male), race (self-identified and categorized as White, Black, Asian, or Mixed/Other), level of education (A-levels or above or less than A-levels), assessment center (Cheadle, Reading, Newcastle, or Bristol), and number of APOE-ε4 alleles. At recruitment, age and sex information were obtained from a central registry and subsequently updated by participants. Participants self-reported their race and education through a touchscreen questionnaire at baseline. The number of APOE-ε4 alleles (0, 1, or 2) was determined by the single nucleotide polymorphisms rs7412 and rs429358.^28^

### Statistical Analyses

We summarized demographic characteristics of the subsamples with MRI data who completed each of the following cognitive outcomes: fluid intelligence, numeric memory, prospective memory, pairs matching, Trail Making A, Trail Making B, reaction time, and symbol digit substitution. To evaluate the associations between regional volumes and cognitive outcomes, we used linear regression for all cognitive outcomes except for prospective memory, for which we used logistic regression (1 for correct on the first attempt and 0 otherwise). All models were adjusted for age, age squared, sex, race, the number of APOE-ε4 alleles, education, and assessment center as a proxy for geographic location. To facilitate comparison of effect sizes across ICV correction approaches, ROIs, and cognitive outcomes, we *z*-standardized all corrected and crude regional volumes, ICV, and continuous cognitive outcomes by subtracting the sample mean and dividing by the sample standard deviation.

The factorial combination of brain volume measure, cognitive outcome, and ICV correction approaches leads to 2,784 distinct estimates. We evaluated the extent to which different ICV correction approaches yield consistent estimates when applied to the same brain volume ROI measure and cognitive outcome by 1) generating pairwise comparisons for correction approaches across regional volumes and cognitive outcomes; 2) summarizing derived statistics from pairwise comparisons; and 3) creating Manhattan plots to show how rates of statistical significance of associations varies across correction approach. Consistency of estimates using different ICV corrections was evaluated using Spearman-rank correlation coefficients for each possible pair of ICV correction methods and across all ROI and cognitive outcome combinations. We additionally generated two measures for the consistency of findings using alternative ICV correction methods: For the first measure, we calculated the proportion of pairs of correction methods without significantly different estimates across all ROIs using a conservative *z-*test.^29^ For the second measure of consistency, we calculated the proportion of pairs of correction approaches for which associations had consistent signs across all brain regions assessed. Specifically, associations were only considered to have opposite signs if both associations were statistically significantly different from the null.

For the residual approach, we additionally evaluated the impact of varying age thresholds to define the normative sample in associations between the eight selected regional volumes linked to dementia and all cognitive outcomes. Specifically, we defined the normative sample using different age cutoffs (<60, <65, <70, <75, <80) and assessed whether the analytical choice of cutoffs affected estimated associations.

We conducted two sensitivity analyses: First, to assess whether the estimates of the associations with cognitive outcomes are affected by observations at the extreme ends of the regional volume distributions, we additionally fit models excluding participants with regional volumes beyond extreme percentiles (1^st^ and 99^th^; **Supplementary Material 3**). Second, we repeated analyses adjusting only for age and age squared to assess whether results are affected by adjustment set chosen. Finally, we additionally examined incident dementia as an outcome (**Supplementary Material 4**).

## Results

Sample sizes across cognitive outcomes ranged from 27,147 to 39,349 (**Table 1**) with very similar demographic compositions. Slightly over half the sample members were female, average age ranged from 64.2 to 64.9 years, nearly all participants were White (ranging from 96.9% to 97.1%), and a majority reported A-level or above education (ranging from 80.7% to 81.5%). Very few individuals had a dementia diagnosis at the MRI visit (n=27) or developed incident dementia in follow-up (n=44).

**Table 1:** Characteristics of analytic sample for each cognitive outcome.

|  | **Fluid intelligence**<br>N=38,607 | **Numeric memory**<br>N=28,793 | **Prospective memory**<br>N=39,330 | **Pairs matching**<br>N=39,349 | **Trail Making A**<br>N=28,861 | **Trail Making B**<br>N=27,417 | **Reaction time**<br>N=39,100 | **Symbol digit substitution**<br>N=28,150 |
| --- | --- | --- | --- | --- | --- | --- | --- | --- |
| **Female, N (%)** | 20,218 (52.4) | 15,073 (52.3) | 20,579 (52.3) | 20,588 (52.3) | 14,692 (52.3) | 14,315 (52.2) | 20,457 (52.3) | 14,731 (52.3) |
| **Age at assessment (years, SD)** | 64.2 (7.6) | 64.9 (7.6) | 64.3 (7.6) | 64.3 (7.6) | 64.9 (7.6) | 64.7 (7.6) | 64.2 (7.6) | 64.9 (7.6) |
| **Race, N (%)** |  |  |  |  |  |  |  |  |
| White | 37,462 (97.0) | 27,939 (97.0) | 38,129 (96.9) | 38,148 (96.9) | 27,272 (97.0) | 26,609 (97.1) | 37,908 (97.0) | 27,313 (97.0) |
| Asian | 527 (1.4) | 395 (1.4) | 560 (1.4) | 560 (1.4) | 379 (1.3) | 373 (1.4) | 554 (1.4) | 383 (1.4) |
| Black | 331 (0.9) | 247 (0.9) | 341 (0.9) | 341 (0.9) | 243 (0.9) | 237 (0.9) | 339 (0.9) | 245 (0.9) |
| Other | 287 (0.7) | 212 (0.7) | 300 (0.8) | 300 (0.8) | 210 (0.7) | 198 (0.7) | 299 (0.8) | 209 (0.7) |
| **Count of APOE-ε4 alleles, N (%)** |  |  |  |  |  |  |  |  |
| 0 | 27,951 (72.4) | 20,829 (72.3) | 28,452 (72.3) | 28,466 (72.3) | 20,322 (72.3) | 19,830 (72.3) | 28,277 (72.3) | 20,364 (72.3) |
| 1 | 9,802 (25.4) | 7,321 (25.4) | 10,007 (25.4) | 10,012 (25.4) | 7,159 (25.5) | 6,978 (25.5) | 9,958 (25.5) | 7,164 (25.4) |
| 2 | 854 (2.2) | 643 (2.2) | 871 (2.2) | 871 (2.2) | 623 (2.2) | 609 (2.2) | 865 (2.2) | 622 (2.2) |
| **Education, N (%)** |  |  |  |  |  |  |  |  |
| A-levels or above | 31,149 (80.7) | 23,403 (81.3) | 31,589 (80.3) | 31,601 (80.3) | 22,807 (81.2) | 22,345 (81.5) | 31,417 (80.4) | 22,875 (81.3) |
| Less than A-levels | 7,458 (19.3) | 5,390 (18.7) | 7,741 (19.7) | 7,748 (19.7) | 5,297 (18.8) | 5,072 (18.5) | 7,683 (19.6) | 5,275 (18.7) |
| **Assessment Center, N (%)** |  |  |  |  |  |  |  |  |
| Reading | 5,788 (15.0) | 5,794 (20.1) | 5,837 (14.8) | 5,837 (14.8) | 5,794 (20.6) | 5,676 (20.7) | 5,820 (14.9) | 5,795 (20.6) |
| Cheadle | 22,843 (59.2) | 13,038 (45.3) | 23,450 (59.6) | 23,469 (59.6) | 12,355 (44.0) | 12,057 (44.0) | 23,267 (59.5) | 12,357 (43.9) |
| Newcastle | 9,928 (25.7) | 9,914 (34.4) | 9,995 (25.4) | 9,995 (25.4) | 9,907 (35.3) | 9,636 (35.1) | 9,965 (25.5) | 9,951 (35.3) |
| Bristol | 48 (0.1) | 47 (0.2) | 48 (0.1) | 48 (0.1) | 48 (0.2) | 48 (0.2) | 48 (0.1) | 47 (0.2) |

Associations of regional brain volumes with cognitive scores are shown in **Figure 1** and **Figure S1**. The associations varied in both sign and magnitude across ICV correction approaches. Crude estimates were typically farthest from the null, and proportional approaches were typically closest to the null, but sometimes with reversed sign compared to other approaches. For example, when using the crude approach, an increase of 0.114 (95% confidence interval [CI] 0.103 to 0.125) in fluid intelligence was associated with each unit increase in hippocampal volume. However, when using the adjustment approach, the increase was 0.055 (95% CI 0.043 to 0.068), while the proportional approach showed a decrease of -0.025 (95% CI -0.035 to -0.014).

**Figure 1:**
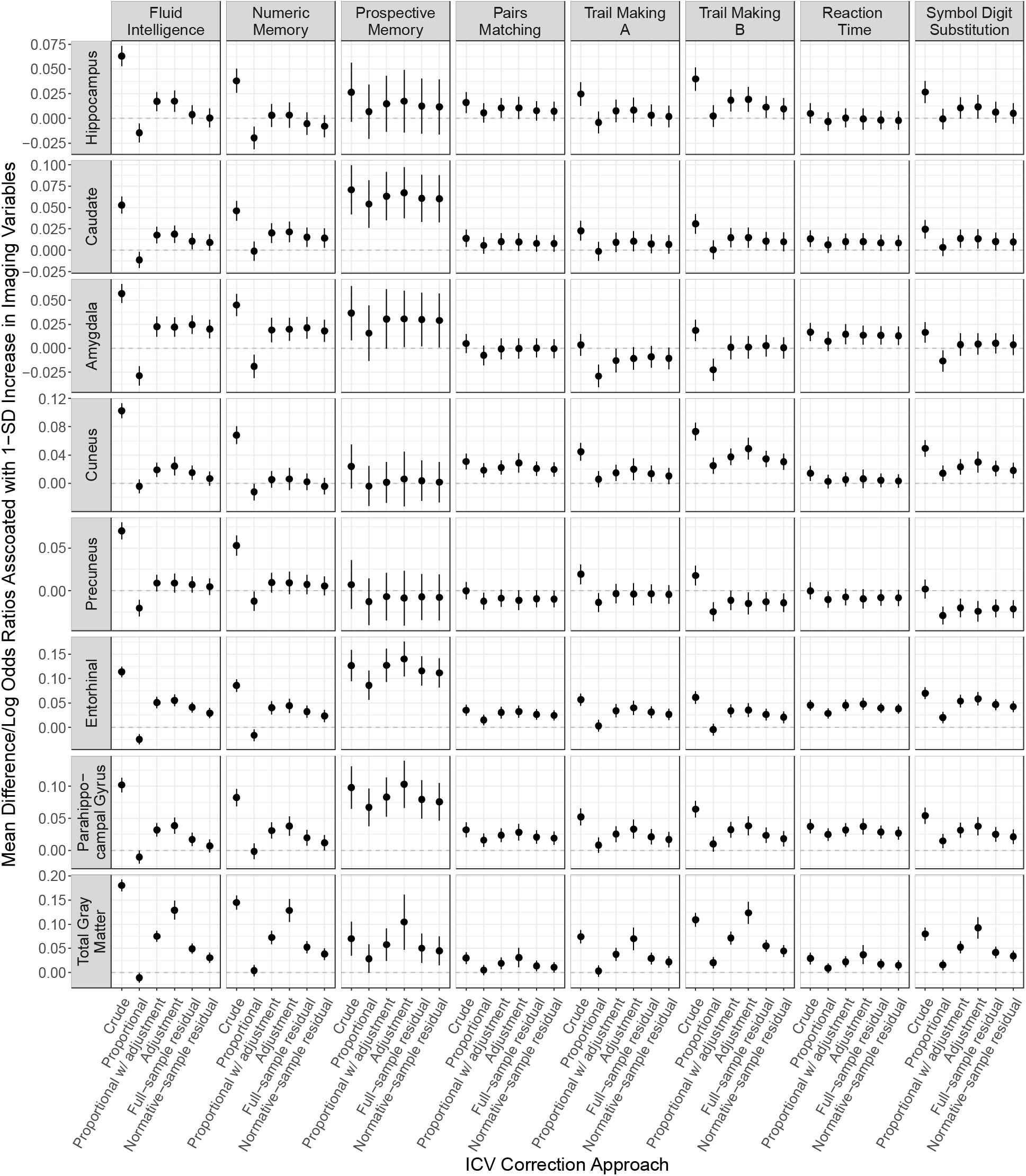
Associations of volumes of the cuneus, entorhinal cortex, parahippocampal gyrus, precuneus, caudate nucleus, hippocampus, amygdala, and total grey matter with each cognitive outcome. Cognitive scores are signed such that higher values indicate better cognition. Specifically, cognitive scores for pairs matching, Trail Making A, Trail Making B, and reaction time refer to negative one times the score on those cognitive tests. Models adjusted for age, age squared, sex, race, the number of APOE-ε4 alleles, education, and assessment center.

**Figure 2** shows comparisons across ICV correction approaches for the associations of all brain regions assessed with fluid intelligence, numeric memory, and Trail Making A and B. Specifically, it shows pairwise scatterplots of estimated associations, as well as Spearman-rank correlations and both measures of consistency for the 58 brain regions evaluated. Estimates when ICV corrections are based on the adjustment approach are highly correlated (*ρ* ≥ 0.89) with coefficients from the residual approach. These two approaches both show only weak correlations with the proportional approach (*ρ* = 0.28 with the adjustment approach and *ρ* = 0.25 with the residual approach, both for fluid intelligence). Different ICV correction approaches sometimes produce both qualitatively different results. The proportional approach was most likely to produce an association with a reversed sign that reaches statistical significance. Even when point estimates are not of the opposite side of the null, estimates may be statistically significantly inconsistent. Supplementary **Figure S2** is the same plot for the remaining cognitive outcomes.

**Figure 2:**
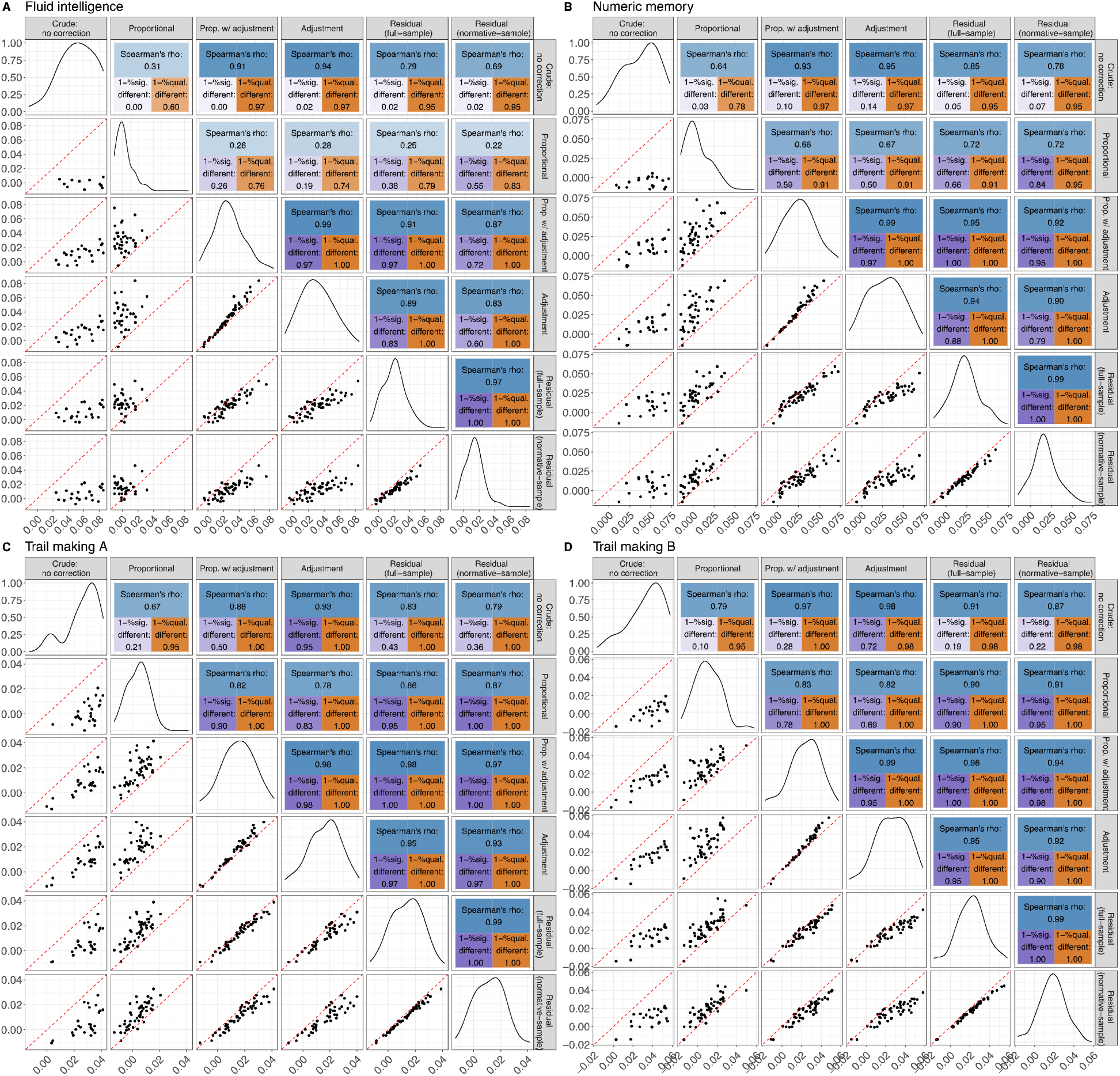
Comparison of coefficients based on alternative ICV corrections for estimated associations of 58 brain region volume measures with A. fluid intelligence, B. numeric memory, C. Trail Making A, and D. Trail Making B. Pairwise comparisons for estimated associations between volumetric measures and fluid intelligence among six total intracranial volume (ICV) correction approaches. Diagonal panels are smoothed density estimates for the distribution of associations across all 58 brain regions assessed under the corresponding ICV correction approach. Lower diagonal panels are scatterplots for the associations under each pair of ICV correction approaches across all 58 brain regions assessed. Upper diagonal panels show the correlation and two consistency measures between each pair. Spearman’s rho (range from -1 to 1) stands for Spearman-rank correlation coefficient; 1 – %sig. different (range from 0 to 1) is one minus the proportion of pairs with significantly different estimates across all ROIs using a conservative Z-test; and 1 – %qual. different (range from 0 to 1) is the one minus proportion of pairs with significantly opposite signs. Color intensity represents the value, with fully saturated colors indicating a 1.

Discrepancies between ICV correction approaches were prominent for fluid intelligence, numeric memory, and Trail Making A and B. Numeric memory and Trail Making A and B are often used in clinical settings as indicators of simple attention/working memory and executive function, respectively, are known to be affected in more advanced stages of Alzheimer’s disease and are prominent areas of cognition affected in other forms of dementia (e.g., frontotemporal dementia).^30–32^ Associations with prospective memory were less appreciably affected by the ICV correction approach, but this may reflect a single limited assessment task in UK Biobank.

**Figure 3** shows brain-wide associations in a Manhattan plot^33^ for all cognitive outcomes and across all brain regions evaluated, comparing ICV correction approaches. Which and how many associations are statistically significant varies with ICV correction approach and cognitive outcome assessed. The crude approach produces the smallest *p*-values, consistent with the premise that failing to correct for ICV inflates associations and statistical significance.

**Figure 3:**
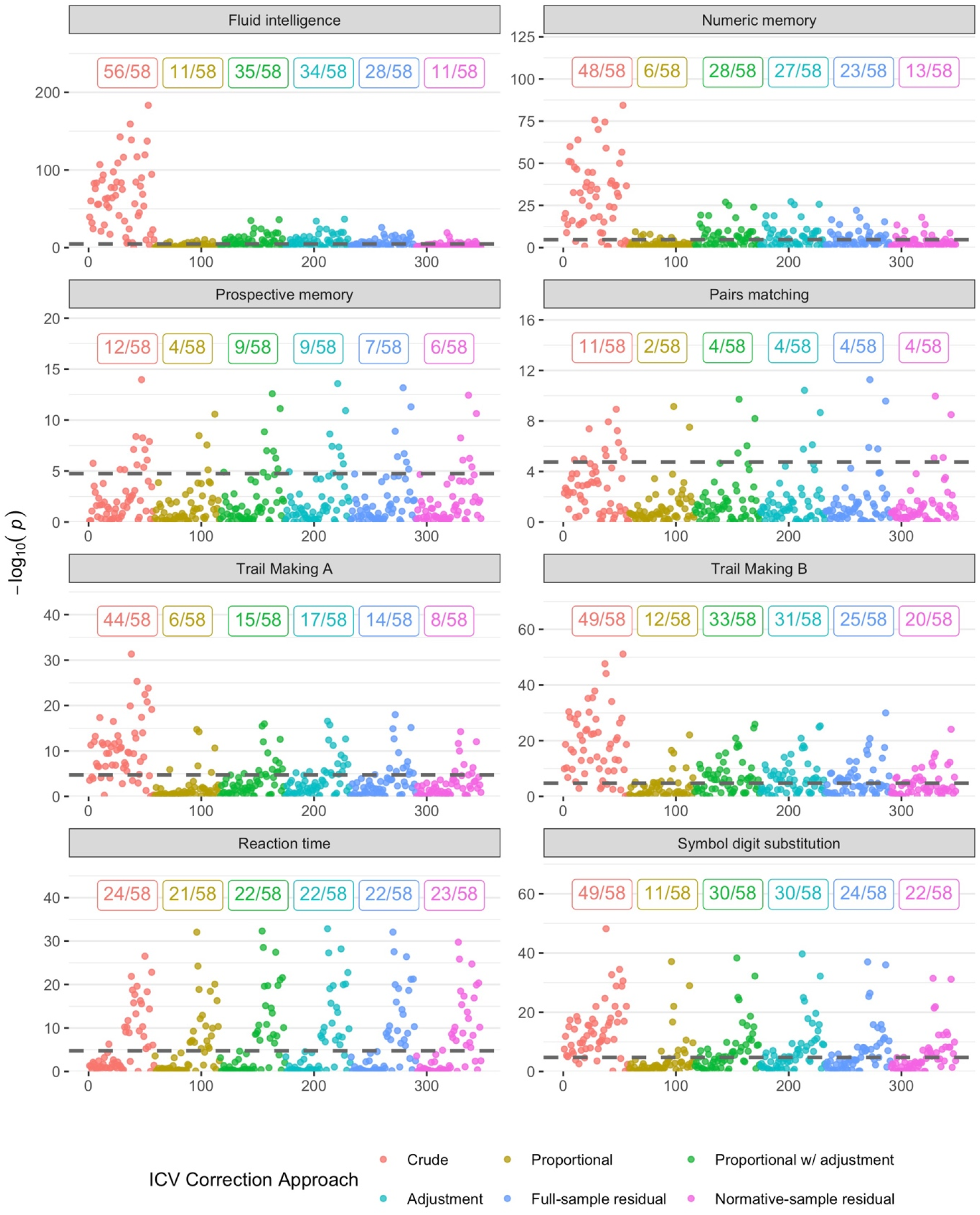
Manhattan plot for association of cognitive outcomes with 58 regional brain volumes, comparing 6 ICV correction approaches. Dashed lines indicate the Bonferroni-corrected threshold for brain-wide significance (p<1.8×10^-5^). Number annotations indicate the number of brain-wide significant regional associations across all 58 brain regions assessed.

The full-sample and normative-sample (age < 60) residual approaches produced highly correlated associations (**Figure 2**), with comparable *p*-values (**Figure 3**). However, estimated associations can vary, as shown in **Figure 1**, with estimates with the normative-sample residual approach typically attenuated relative to those of approach the full-sample residual approach. **Figure 4** extends this, showing how using an increasingly older, and presumably less healthy sample with more age-associated neurodegeneration, delivers larger estimated associations between hippocampal volume and fluid intelligence. Consistent with results in **Figure 1**, **Figure 4** shows an increasingly younger sample produces an attenuated association between selected regions and fluid intelligence, numeric memory, and Trail Making B. This highlights that definition of normative sample can affect estimated associations. In sensitivity analyses, we find that results are comparable excluding outliers in ICV and with different adjustment sets. See supplemental **Figures S3**-**S6**. In addition, we did not observe similar inconsistency in time-to-event analysis (**Figure S7**).

**Figure 4:**
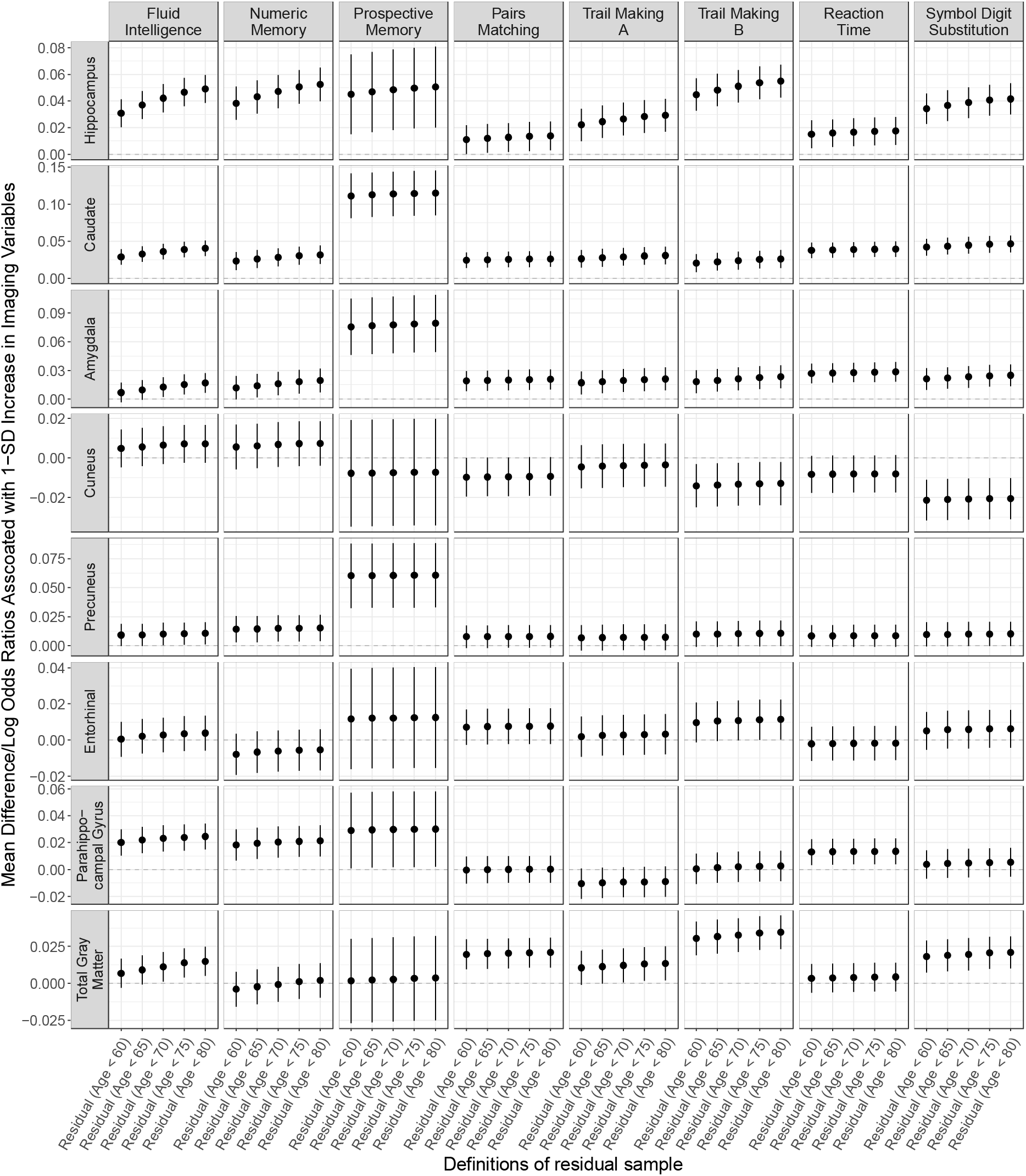
Effect of definition of residual sample on associations between volumes of the cuneus, entorhinal cortex, parahippocampal gyrus, precuneus, caudate nucleus, hippocampus, amygdala, and total grey matter and all cognitive outcomes.

## Discussion

We evaluated whether estimated associations between regional brain volumetric measures and cognitive outcomes differed when using crude volumes or five alternative approaches to correct for ICV using the large UK Biobank MRI subsample. Although estimates based on the adjustment and residual approaches were similar, estimates based on the proportional and crude approaches were inconsistent. Inconsistencies were largest when estimates from the adjustment and residual approaches were further from the null.

The proportional, adjustment, and residual approaches are all commonly used strategies^10^ to correct for ICV, and crude estimates are frequently presented as well.^13^ However, we found that the proportional approach frequently produced estimates that were inconsistent with understanding of neurobiology and even have the opposite sign of estimates derived with adjustment and residual approaches. As an example, the hippocampus plays a vital role in encoding and consolidation of new memories.^34^ Numerous studies have demonstrated an association between hippocampal atrophy and poorer neuropsychological test performance, particularly with regards to memory tasks.^35–38^ This relationship is further corroborated by existing experimental studies and clinical case studies involving direct damage to the hippocampus ^39–41^. However, estimates for the association between hippocampal volume and cognitive outcomes were inconsistent across ICV correction approaches: the proportional indicated that larger hippocampal volume was associated with worse cognition, which is contravenes extant understanding of neurobiology and neurodegenerative diseases (see **Figure 1**).^37,42^ While more often consistent, even adjustment and residual approaches do not always produce consistent estimated associations, particularly since the associations produced with the residual approach depended on the normative sample adopted (see **Figure 4**). Specifically, we find using increasingly younger normative samples attenuated estimated associations between hippocampal volume and fluid intelligence.

These results have important implications for reproducibility in neuroimaging studies, our understanding of disease biology, and intervention evaluation. We recognize that these ICV correction methods represent different biological constructs and that method employed may depend on the nature of the research question. For example, in some studies, we may not want to account for ICV to account for perinatal and childhood factors that influence cranium size and regional volumes. However, findings from different studies may diverge merely because of the selected ICV correction approach—ostensibly a minor statistical decision. Estimated associations from crude volumes tended to be further from the null than findings after ICV correction, suggesting that ICV captures confounding by lifetime peak brain size or childhood growth. If atrophy-related neurodegeneration is the construct we intend to capture, ICV correction will often be necessary. Moreover, it is important to note that ICV correction is also crucial for accurately identifying and interpreting volumetric measures that are associated with cognitive functioning or dementia symptoms.^5^ Our findings suggest the proportional approach may be misleading and, if used, should be interpreted in conjunction with results from other approaches. The adjustment and residual approaches tend to produce comparable estimates and it may be reasonable to favor these two approaches. However, estimated associations with residual approaches can vary with reference sample used (**Figure 4**). Even more concerning, as the age of the normative sample is decreased, biologically plausible effects are increasingly attenuated, approaching the null as the sample becomes younger and healthier. In addition, the residual approach, when not applied to a separate normative sample, ideally would include a standard error correction to account for the fact that the two-stages of estimation are performed on overlapping samples. Thus, if ICV is to be corrected for, we would tend to favor adjustment over the other methods since it is the simplest to implement while producing biologically plausible associations.

Our results are consistent with small prior studies indicating that ICV correction approach can affect study results. Previous studies have examined whether associations of sex and age with volumetric measure persisted across ICV correction approaches.^14,15,43^ Prior work has also examined associations between brain volumetric measures and cognition,^27^ finding that ICV correction approach altered associations, with flipped direction of association for the proportional method. This study was performed in two smaller and select samples (*N* = 406 and 724) and examined only the first item of word recall from the ADAS-Cog. The small samples have left substantial uncertainty in whether their conclusions hold for larger and less select samples. For example, previous work found that the association between hippocampal volumes and cognition was not statistically significantly different across ICV correction approach, possibly due to imprecise estimates. Our sample, with 25 to 35 times as many participants as the previous study and a much broader range of volumetric measures and cognitive outcomes assessed,^27^ provides far more conclusive findings on the importance of ICV correction approach.

Our study has several strengths in addition to the large sample: these include comprehensive evaluation of ICV correction approaches and a wide range of cognitive outcomes evaluated. Employing a BWAS-inspired approach, we examined a large combination of associations between cognitive tests and regional brain volumes for a total of 2,784 regressions with several measures of consistency to further support the robustness of our findings. Our study has several limitations. First, our results only pertained to cross-sectional evaluation of MRI volumetric measures and cognitive outcomes. Other neuroimaging measures—including cortical thicknesses, diffusion tensor imaging, and longitudinal change in volumetric measures^44^—may warrant further investigation.^3^ UK Biobank participants are known to be healthier on average than the general UK population, have higher socioeconomic status, and are predominately White.^45^ Patterns may differ in more diverse populations.^46^ Finally, did not evaluate alternative ICV estimation methods,^47^ and we did not examine less commonly used methods (e.g., weighted ICV matching^16^).

In conclusion, different ICV correction approaches can produce substantively different estimated associations between MRI-derived measures of brain volume and cognitive outcomes. These differences are largest when the associations are large. The proportional approach is most likely to produce estimates that are inconsistent with adjustment or residual control approaches and biologically implausible. Residual and adjustment approaches are more plausible but since they may produce different results, results based on only one of the approaches should be interpreted with caution.

## Data Availability

All data produced in the present study are available upon reasonable request to the authors.

