## Supplementary Material 1-4 and Supplementary Figure 1-7 for "Comparison of approaches to control for intracranial volume in research on the association of brain volumes with cognitive outcomes"

**Supplementary Material 1. UK Biobank field IDs for MRI variables and covariates.**

| **Variable** | **UK Biobank Field ID** | |
| --- | --- | --- |
| ***MRI variables (cortical)*** | **Left hemisphere** | **Right hemisphere** |
|  | **Total brain** | |
| Volume of caudal anterior cingulate | 27205 | 27298 |
| Volume of caudal middle frontal | 27206 | 27299 |
| Volume of cuneus | 27207 | 27300 |
| Volume of entorhinal | 27208 | 27301 |
| Volume of fusiform | 27209 | 27302 |
| Volume of inferior parietal | 27210 | 27303 |
| Volume of inferior temporal | 27211 | 27304 |
| Volume of isthmus cingulate | 27212 | 27305 |
| Volume of lateral occipital | 27213 | 27306 |
| Volume of lateral orbitofrontal | 27214 | 27307 |
| Volume of lingual | 27215 | 27308 |
| Volume of medial orbitofrontal | 27216 | 27309 |
| Volume of middle temporal | 27217 | 27310 |
| Volume of parahippocampal gyrus | 27218 | 27311 |
| Volume of paracentral | 27219 | 27312 |
| Volume of parsopercularis | 27220 | 27313 |
| Volume of parsorbitalis | 27221 | 27314 |
| Volume of parstriangularis | 27222 | 27315 |
| Volume of pericalcarine | 27223 | 27316 |
| Volume of postcentral | 27224 | 27317 |
| Volume of posterior cingulate | 27225 | 27318 |
| Volume of precentral | 27226 | 27319 |
| Volume of precuneus | 27227 | 27320 |
| Volume of rostral anterior cingulate | 27228 | 27321 |
| Volume of rostral middle frontal | 27229 | 27322 |
| Volume of superior frontal | 27230 | 27323 |
| Volume of superior parietal | 27231 | 27324 |
| Volume of superior temporal | 27232 | 27325 |
| Volume of supramarginal | 27233 | 27326 |
| Volume of transverse temporal | 27234 | 27327 |
| Volume of insula | 27235 | 27328 |
| ***MRI variables (subcortical)*** | **Left hemisphere** | **Right hemisphere** |
|  | **Total brain** | |
| Volume of posterior corpus callosum | 26531 | |
| Volume of mid-posterior corpus callosum | 26532 | |
| Volume of central corpus callosum | 26533 | |
| Volume of mid-anterior corpus callosum | 26534 | |
| Volume of anterior corpus callosum | 26535 | |
| Volume of cortex | 26552 | 26583 |
| Volume of cerebral white matter | 26553 | 26584 |
| Volume of lateral ventricle | 26554 | 26585 |
| Volume of inferior lateral ventricles | 26555 | 26586 |
| Volume of cerebellum white matter | 26556 | 26587 |
| Volume of cerebellum cortex | 26557 | 26588 |
| Volume of thalamus proper | 26558 | 26589 |
| Volume of caudate | 26559 | 26590 |
| Volume of putamen | 26560 | 26591 |
| Volume of pallidum | 26561 | 26592 |
| Volume of hippocampus | 26562 | 26593 |
| Volume of amygdala | 26563 | 26594 |
| Volume of accumbens area | 26564 | 26595 |
| Volume of ventral DC | 26565 | 26596 |
| Volume of choroid-plexus | 26567 | 26598 |
| Volume of subcortical grey matter | 26517 | |
| Volume of total grey matter | 26518 | |
| Volume of 3rd ventricle | 26523 | |
| Volume of 4th ventricle | 26524 | |
| Volume of brain stem | 26526 | |
| Volume of cerebrospinal fluid | 26527 | |
| Volume of optic chiasm | 26530 | |
| ***Covariates*** | | |
| Age | 21022 | |
| Sex | 31 | |
| Race | 21000 | |
| Education | 6138 | |
| Assessment center | 54 | |

**Supplementary Material 2: UK Biobank cognitive tests**

**Fluid Intelligence (**[**Category 100027**](https://biobank.ndph.ox.ac.uk/showcase/label.cgi?id=100027)**,** [**Field ID 20016**](https://biobank.ndph.ox.ac.uk/showcase/field.cgi?id=20016)**)**

The capacity to solve problems that require logic and reasoning ability, independent of acquired knowledge. Participants had 2 minutes to complete as many questions as possible from the test. The score is a simple unweighted sum of the number of correct answers given to the 13 fluid intelligence questions. Participants who did not answer all of the questions within the allotted 2-minute limit are scored as zero for each of the unattempted questions.

**Numeric Memory (**[**Category 100029**](https://biobank.ndph.ox.ac.uk/showcase/label.cgi?id=100029)**,** [**Field ID 4282**](https://biobank.ndph.ox.ac.uk/showcase/field.cgi?id=4282)**)**

This test is designed to assess numeric short-term memory. Participants are shown a 2-digit number to remember. The number then disappeared and after a short while they were asked to enter the number onto the screen. The number became one digit longer each time they remembered correctly (up to a maximum of 12 digits). The score is longest number correctly recalled during the numeric memory test.

**Prospective Memory (**[**Category 100031**](https://biobank.ndph.ox.ac.uk/showcase/label.cgi?id=100031), [**Field ID 20018**](https://biobank.ndph.ox.ac.uk/showcase/field.cgi?id=20018)**)**

Early in the cognitive section, the participant is shown the message "At the end of the games we will show you four coloured shapes and ask you to touch the Blue Square. However, to test your memory, we want you to actually touch the Orange Circle instead." Scores were considered correct only if correct on first attempt.

**Pairs Matching (**[**Category 100030**](https://biobank.ndph.ox.ac.uk/showcase/label.cgi?id=100030)**,** [**Field ID 399**](https://biobank.ndph.ox.ac.uk/showcase/field.cgi?id=399)**)**

Participants are asked to memorize the position of as many matching pairs of cards as possible. The cards are then turned face down on the screen and the participant is asked to touch as many pairs as possible in the fewest tries. Multiple rounds were conducted. The first round used 3 pairs of cards and the second 6 pairs of cards. The score is the negative of the number of mistakes, with the highest score being zero (no mistakes).

**Trail Making A and B (**[**Category 505**](https://biobank.ndph.ox.ac.uk/showcase/label.cgi?id=505)**,** [**Field IDs 6348**](https://biobank.ndph.ox.ac.uk/showcase/field.cgi?id=6348)**,** [**6350**](https://biobank.ndph.ox.ac.uk/showcase/field.cgi?id=6350)**)**

Participants were presented with sets of digits/letters in circles scattered around the screen and asked to click on them sequentially according to a specific algorithm. The score is the negative of the amount of time it took to complete the test.

**Reaction Time (**[**Category 100032**](https://biobank.ndph.ox.ac.uk/showcase/label.cgi?id=100032)**,** [**Field ID 20023**](https://biobank.ndph.ox.ac.uk/showcase/field.cgi?id=20023)**)**

The participant is shown two cards at a time; if both cards are the same, they press a button-box that is on the table in front of them as quickly as possible. The score is the negative of the amount of time it took to complete the test.

**Symbol Digit Substitution (**[**Category 502**](https://biobank.ndph.ox.ac.uk/showcase/label.cgi?id=502), [**Field ID 6325**](https://biobank.ndph.ox.ac.uk/showcase/field.cgi?id=6325)**)**

Participant are presented with one grid linking symbols to single-digit integers and a second grid containing only the symbols. They are then asked to indicate the numbers attached to each of the symbols in the second grid using the first one as a key. The score is the number of symbols correctly matched to digits.

**Supplementary Material 3: Outlier detection and exclusion for regional volumes.**

In the sensitivity analysis, the primary analyses were repeated removing the extreme (top and bottom 1%) values of the regional volumes. For each region of interest (ROI), we detected the extreme values for crude volume, proportional volume, and residual volumes from two residual approaches. Then we excluded participants with at least one extreme value. In addition, we also excluded participants with extreme intracranial volume (ICV). The numbers of excluded participants for each ROI and cognitive outcome are listed in the table below.

| **ROI** | **Fluid intelligence** | **Numeric memory** | **Prospective memory** | **Pairs matching** | **Trail Making A** | **Trail Making B** | **Reaction time** | **Symbol digit substitution** |
| --- | --- | --- | --- | --- | --- | --- | --- | --- |
| Volume of caudal anterior cingulate | 1,874 | 1,439 | 1,914 | 1,915 | 1,422 | 1,394 | 1,901 | 1,422 |
| Volume of caudal middle frontal | 1,952 | 1,466 | 1,988 | 1,988 | 1,442 | 1,411 | 1,977 | 1,440 |
| Volume of cuneus | 1,958 | 1,476 | 2,002 | 2,002 | 1,459 | 1,418 | 1,992 | 1,454 |
| Volume of entorhinal | 1,771 | 1,332 | 1,824 | 1,824 | 1,308 | 1,270 | 1,808 | 1,311 |
| Volume of fusiform | 1,997 | 1,494 | 2,050 | 2,051 | 1,472 | 1,437 | 2,041 | 1,470 |
| Volume of inferior parietal | 1,981 | 1,457 | 2,028 | 2,028 | 1,431 | 1,395 | 2,014 | 1,433 |
| Volume of inferior temporal | 1,970 | 1,484 | 2,026 | 2,027 | 1,446 | 1,401 | 2,015 | 1,446 |
| Volume of isthmus cingulate | 1,947 | 1,472 | 1,989 | 1,989 | 1,448 | 1,412 | 1,982 | 1,446 |
| Volume of lateral occipital | 1,981 | 1,472 | 2,029 | 2,029 | 1,445 | 1,407 | 2,021 | 1,437 |
| Volume of lateral orbitofrontal | 2,012 | 1,476 | 2,064 | 2,064 | 1,456 | 1,422 | 2,048 | 1,449 |
| Volume of lingual | 1,953 | 1,494 | 1,987 | 1,988 | 1,473 | 1,434 | 1,974 | 1,465 |
| Volume of medial orbitofrontal | 2,028 | 1,403 | 2,079 | 2,080 | 1,368 | 1,325 | 2,067 | 1,363 |
| Volume of middle temporal | 2,021 | 1,501 | 2,070 | 2,071 | 1,468 | 1,427 | 2,053 | 1,466 |
| Volume of parahippocampal gyrus | 1,926 | 1,456 | 1,978 | 1,978 | 1,439 | 1,385 | 1,963 | 1,440 |
| Volume of paracentral | 1,969 | 1,507 | 2,015 | 2,016 | 1,483 | 1,440 | 2,003 | 1,483 |
| Volume of parsopercularis | 1,919 | 1,402 | 1,966 | 1,966 | 1,372 | 1,338 | 1,955 | 1,376 |
| Volume of parsorbitalis | 2,000 | 1,470 | 2,047 | 2,047 | 1,454 | 1,419 | 2,035 | 1,454 |
| Volume of parstriangularis | 1,959 | 1,433 | 2,002 | 2,003 | 1,405 | 1,369 | 1,993 | 1,398 |
| Volume of pericalcarine | 1,986 | 1,483 | 2,023 | 2,024 | 1,456 | 1,422 | 2,020 | 1,453 |
| Volume of postcentral | 2,016 | 1,484 | 2,066 | 2,067 | 1,462 | 1,412 | 2,056 | 1,465 |
| Volume of posterior cingulate | 1,899 | 1,434 | 1,947 | 1,947 | 1,422 | 1,384 | 1,935 | 1,417 |
| Volume of precentral | 1,991 | 1,506 | 2,039 | 2,039 | 1,485 | 1,442 | 2,026 | 1,486 |
| Volume of precuneus | 2,001 | 1,489 | 2,044 | 2,045 | 1,462 | 1,417 | 2,031 | 1,458 |
| Volume of rostral anterior cingulate | 1,984 | 1,467 | 2,035 | 2,035 | 1,441 | 1,403 | 2,021 | 1,449 |
| Volume of rostral middle frontal | 1,990 | 1,481 | 2,039 | 2,039 | 1,455 | 1,412 | 2,021 | 1,451 |
| Volume of superior frontal | 1,995 | 1,481 | 2,049 | 2,049 | 1,461 | 1,418 | 2,030 | 1,456 |
| Volume of superior parietal | 1,983 | 1,499 | 2,035 | 2,036 | 1,472 | 1,432 | 2,022 | 1,471 |
| Volume of superior temporal | 2,017 | 1,473 | 2,080 | 2,080 | 1,437 | 1,393 | 2,064 | 1,435 |
| Volume of supramarginal | 1,951 | 1,446 | 1,991 | 1,992 | 1,411 | 1,381 | 1,981 | 1,413 |
| Volume of transverse temporal | 1,930 | 1,401 | 1,981 | 1,981 | 1,388 | 1,345 | 1,970 | 1,388 |
| Volume of insula | 2,022 | 1,496 | 2,080 | 2,081 | 1,476 | 1,433 | 2,066 | 1,472 |
| Volume of posterior corpus callosum | 1,799 | 1,348 | 1,835 | 1,836 | 1,330 | 1,293 | 1,829 | 1,330 |
| Volume of mid-posterior corpus callosum | 1,773 | 1,375 | 1,815 | 1,817 | 1,353 | 1,311 | 1,808 | 1,354 |
| Volume of central corpus callosum | 1,829 | 1,401 | 1,872 | 1,873 | 1,388 | 1,345 | 1,861 | 1,388 |
| Volume of mid-anterior corpus callosum | 1,790 | 1,373 | 1,834 | 1,835 | 1,351 | 1,309 | 1,824 | 1,351 |
| Volume of anterior corpus callosum | 1,871 | 1,440 | 1,917 | 1,918 | 1,414 | 1,386 | 1,909 | 1,419 |
| Volume of cortex | 2,029 | 1,517 | 2,079 | 2,079 | 1,490 | 1,440 | 2,063 | 1,482 |
| Volume of cerebral white matter | 1,958 | 1,494 | 2,000 | 2,000 | 1,460 | 1,425 | 1,987 | 1,466 |
| Volume of lateral ventricle | 1,933 | 1,439 | 1,973 | 1,973 | 1,408 | 1,377 | 1,962 | 1,415 |
| Volume of inferior lateral ventricles | 1,850 | 1,429 | 1,889 | 1,889 | 1,402 | 1,366 | 1,877 | 1,412 |
| Volume of cerebellum white matter | 1,879 | 1,418 | 1,930 | 1,930 | 1,385 | 1,342 | 1,920 | 1,394 |
| Volume of cerebellum cortex | 2,042 | 1,555 | 2,090 | 2,090 | 1,532 | 1,485 | 2,079 | 1,540 |
| Volume of thalamus proper | 2,016 | 1,522 | 2,065 | 2,065 | 1,486 | 1,447 | 2,055 | 1,494 |
| Volume of caudate | 1,924 | 1,482 | 1,976 | 1,978 | 1,461 | 1,417 | 1,964 | 1,461 |
| Volume of putamen | 1,923 | 1,493 | 1,977 | 1,977 | 1,453 | 1,406 | 1,964 | 1,451 |
| Volume of pallidum | 1,971 | 1,488 | 2,028 | 2,028 | 1,457 | 1,412 | 2,013 | 1,460 |
| Volume of hippocampus | 2,013 | 1,551 | 2,070 | 2,070 | 1,512 | 1,468 | 2,058 | 1,519 |
| Volume of amygdala | 1,922 | 1,439 | 1,975 | 1,976 | 1,407 | 1,363 | 1,960 | 1,406 |
| Volume of accumbens area | 1,878 | 1,442 | 1,932 | 1,933 | 1,422 | 1,368 | 1,909 | 1,424 |
| Volume of ventral DC | 2,044 | 1,545 | 2,098 | 2,098 | 1,512 | 1,460 | 2,081 | 1,518 |
| Volume of choroid-plexus | 1,929 | 1,440 | 1,967 | 1,967 | 1,428 | 1,401 | 1,954 | 1,427 |
| Volume of subcortical grey matter | 2,087 | 1,591 | 2,147 | 2,147 | 1,553 | 1,511 | 2,132 | 1,557 |
| Volume of total grey matter | 2,048 | 1,540 | 2,103 | 2,103 | 1,514 | 1,464 | 2,086 | 1,512 |
| Volume of 3rd ventricle | 1,948 | 1,487 | 1,988 | 1,988 | 1,462 | 1,432 | 1,978 | 1,469 |
| Volume of 4th ventricle | 1,882 | 1,404 | 1,918 | 1,919 | 1,381 | 1,357 | 1,913 | 1,380 |
| Volume of brain stem | 2,022 | 1,543 | 2,062 | 2,062 | 1,521 | 1,486 | 2,054 | 1,520 |
| Volume of cerebrospinal fluid | 1,879 | 1,407 | 1,918 | 1,918 | 1,384 | 1,358 | 1,903 | 1,387 |
| Volume of optic chiasm | 1,785 | 1,373 | 1,825 | 1,825 | 1,354 | 1,324 | 1,812 | 1,356 |

**Supplementary Material 4: Time-to-event analysis to examine associations between regional volumes and incident dementia.**

In the time-to-event analysis, we examined the associations between regional volumes and incident dementia under different ICV correction approaches. Specifically, we used Cox proportional hazards regression models to estimate adjusted hazard ratios (HR) between each regional volume and incident all-cause dementia.

We considered the MRI visit as baseline and only included participants with no dementia diagnosis at the MRI visit. Participants were followed up to the first date of all-cause dementia diagnosis, death, or censoring on September 30, 2021 (the latest date of all-cause dementia diagnosis in the MRI subsample), whichever came first. We adjusted for age and sex in the Cox proportional hazards regression models.

**Supplementary Figure S1: Associations between volumes of all regions and all cognitive scores, except for cuneus, entorhinal cortex, parahippocampal gyrus, precuneus, caudate nucleus, hippocampus, amygdala, and total grey matter.**
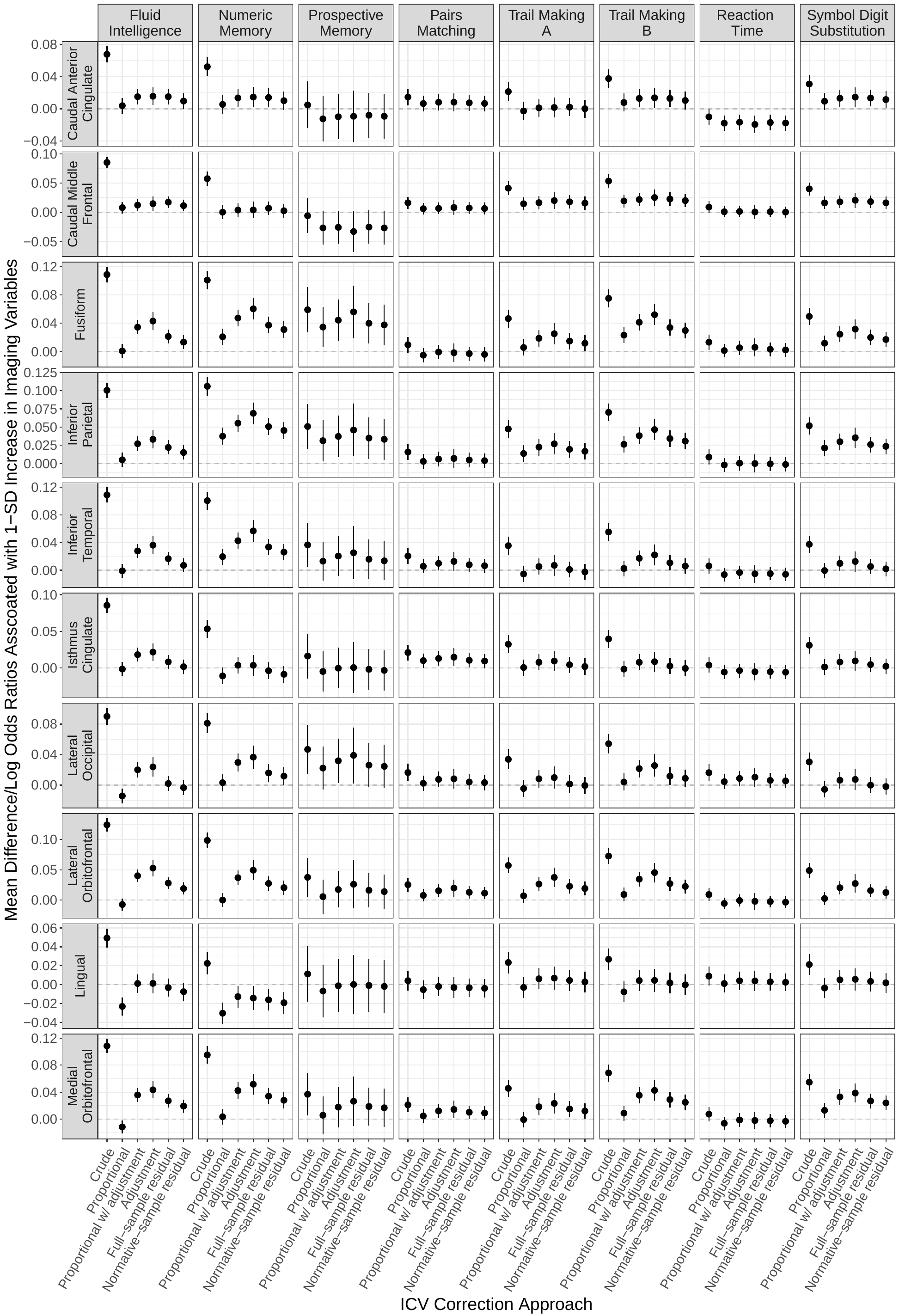

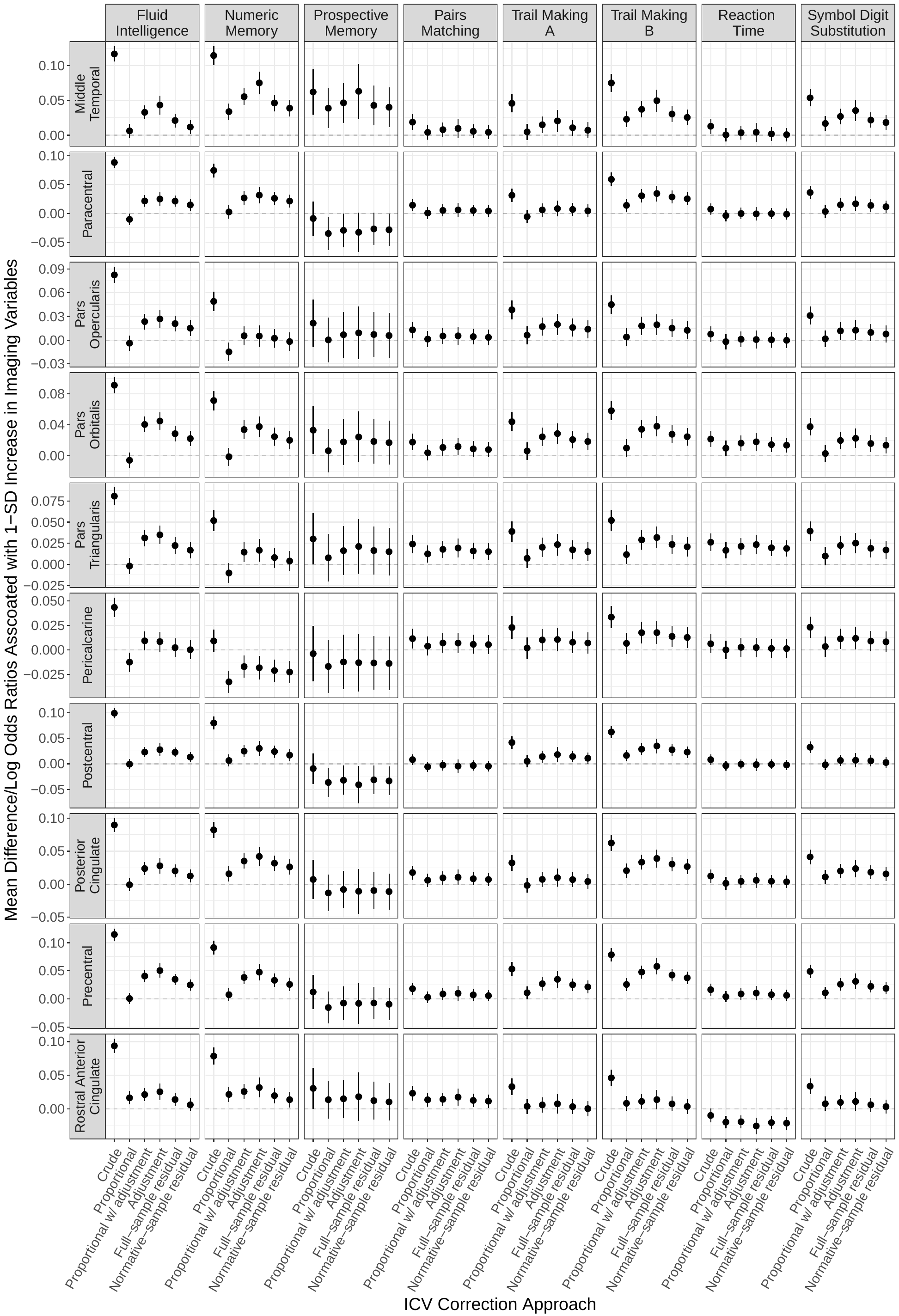

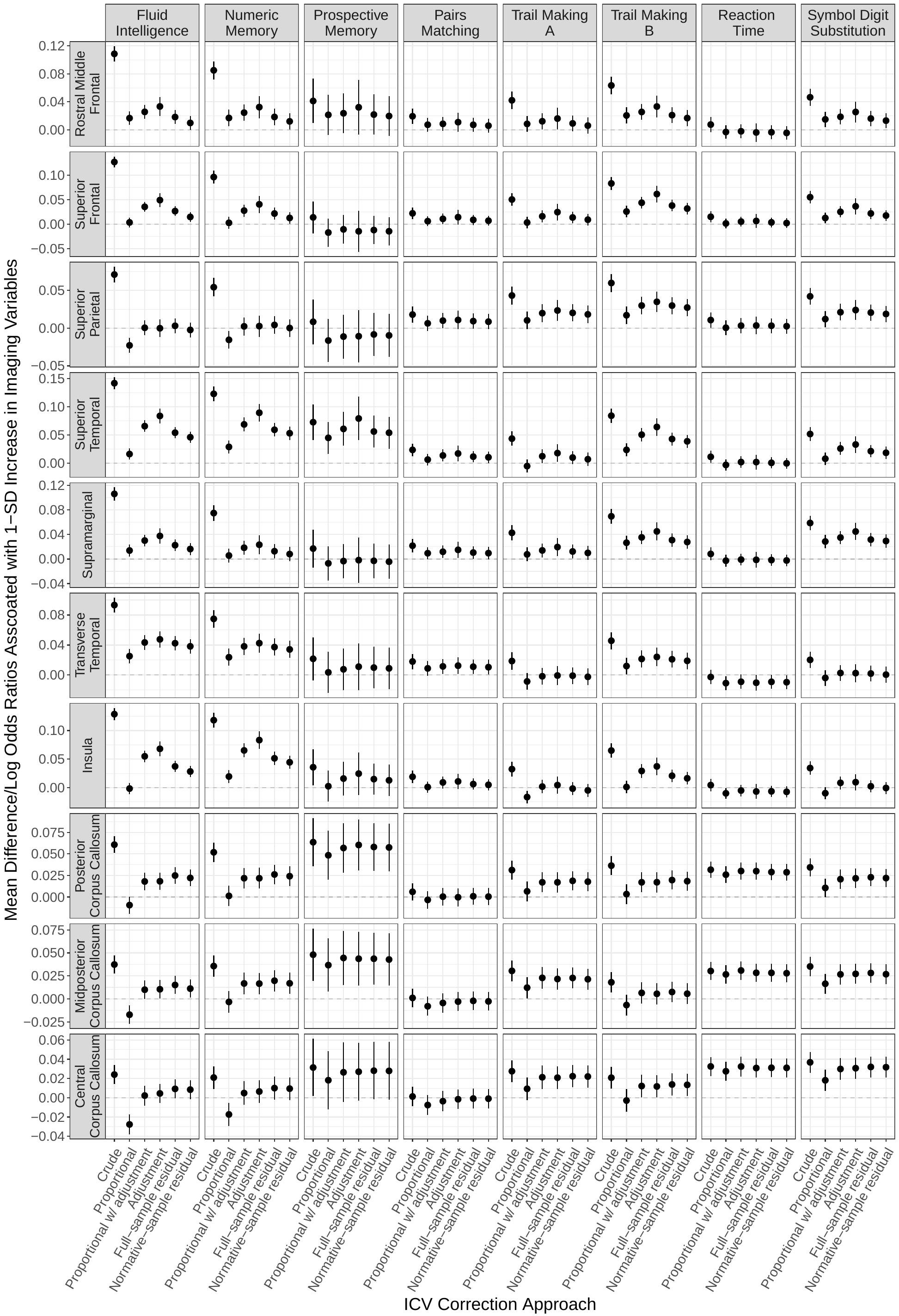

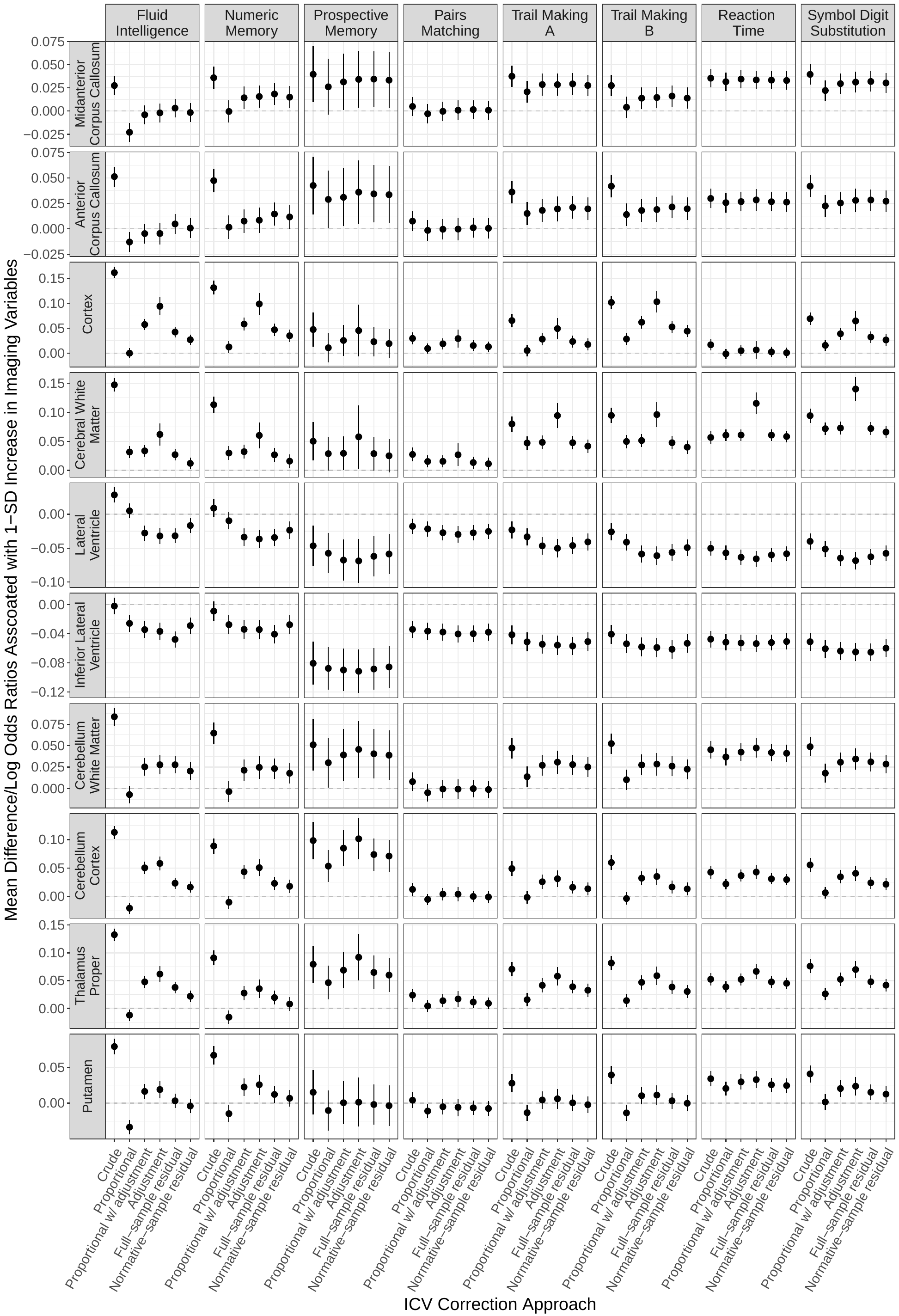

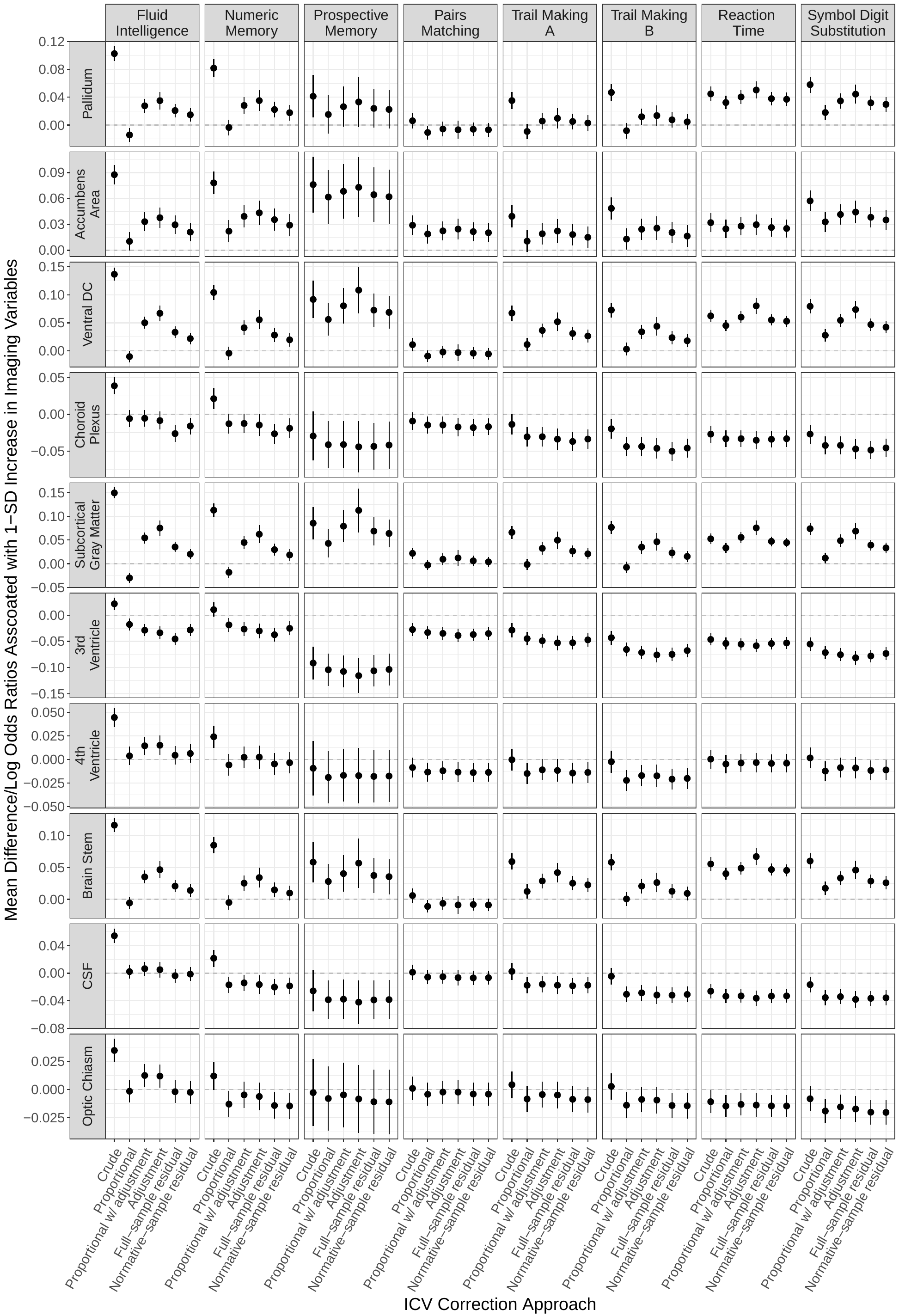

**Supplementary Figure S2: Pairwise correlations and consistencies for estimated associations between volumetric measures and** **A. prospective memory, B. pairs matching, C. reaction time, and D. symbol digit substitution across all 58 brain regions assessed.**

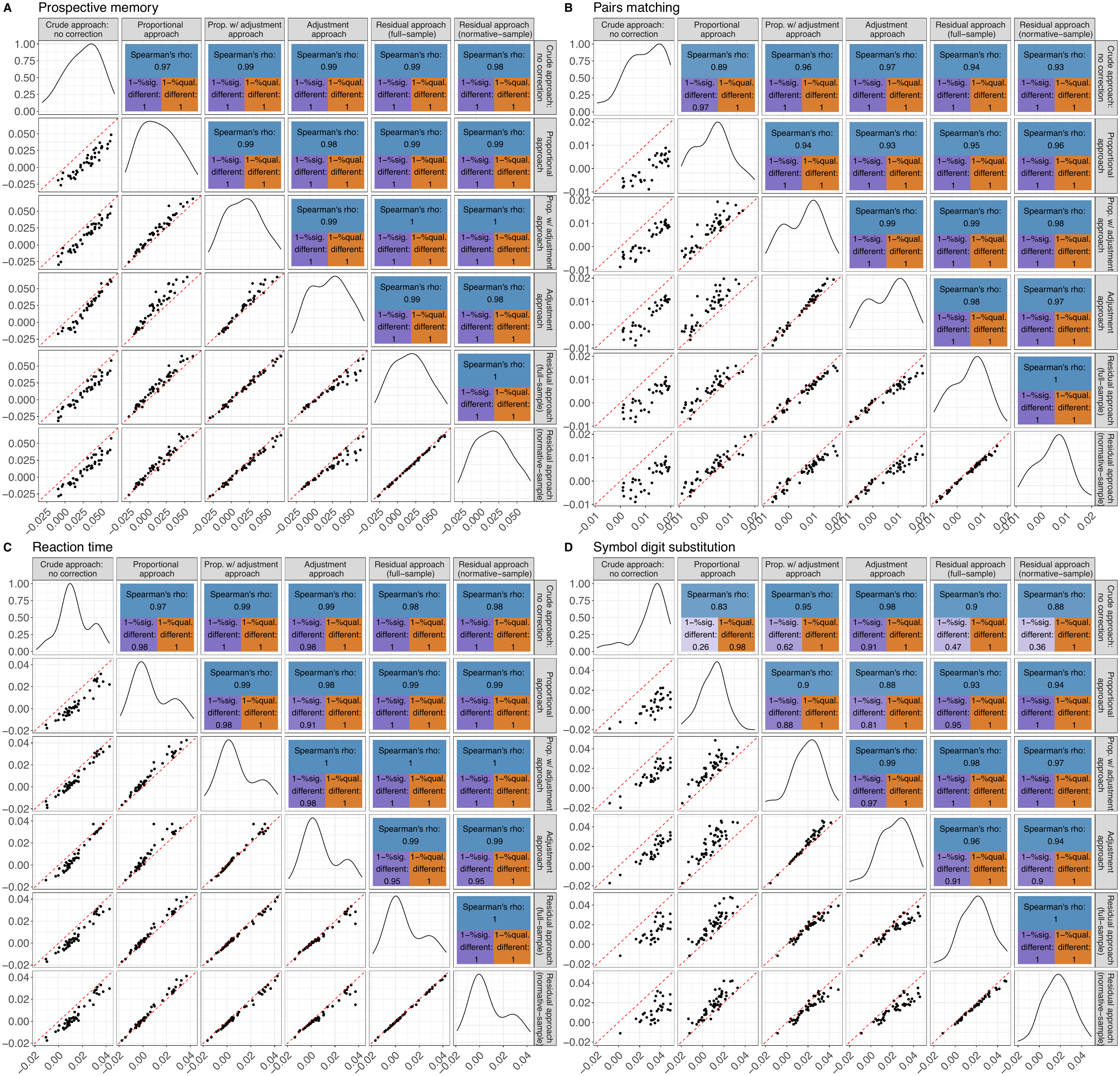

**Supplementary Figure S3: Associations between volumes of all regions and all cognitive scores, excluding outliers of the regional volumes.**
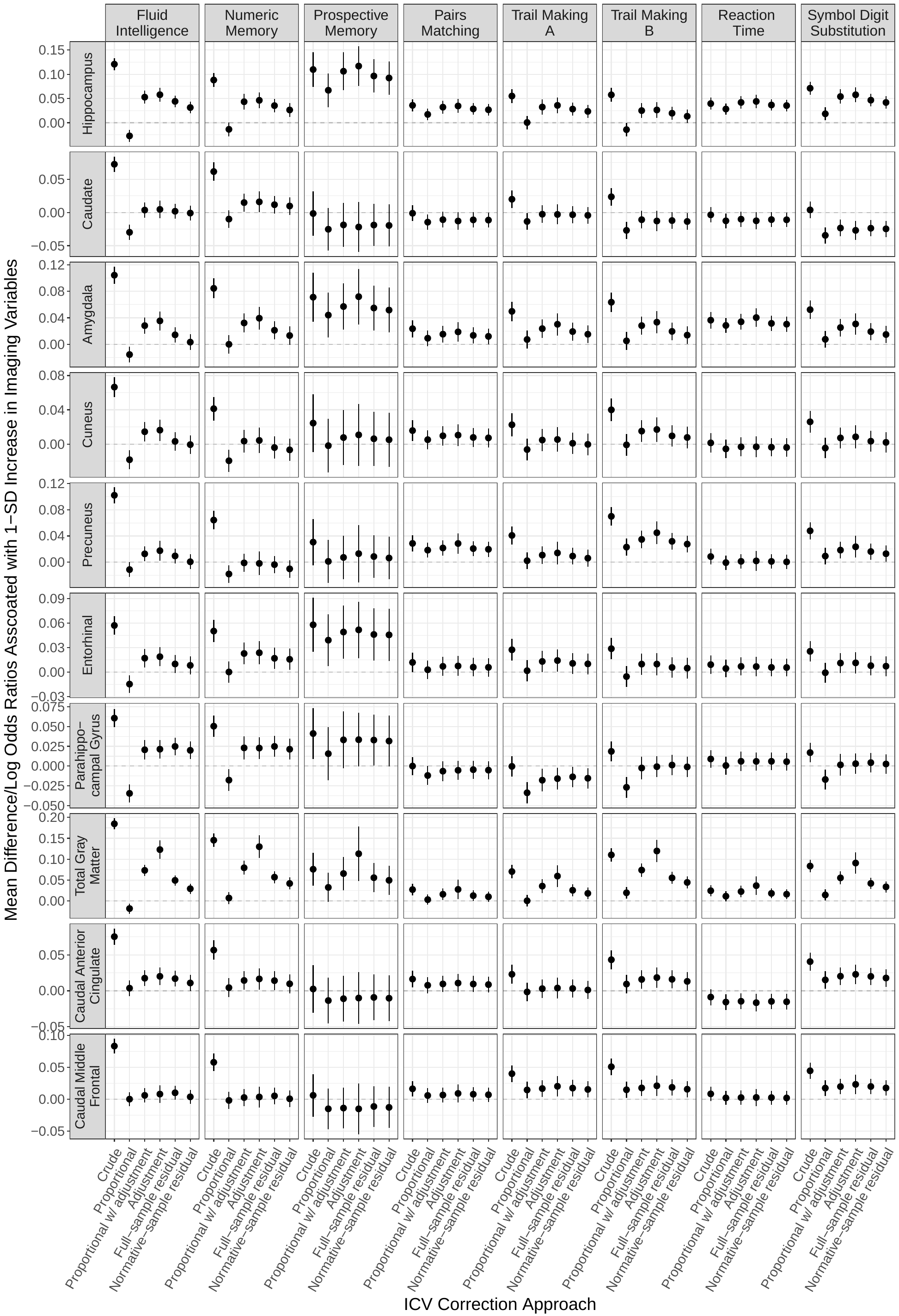

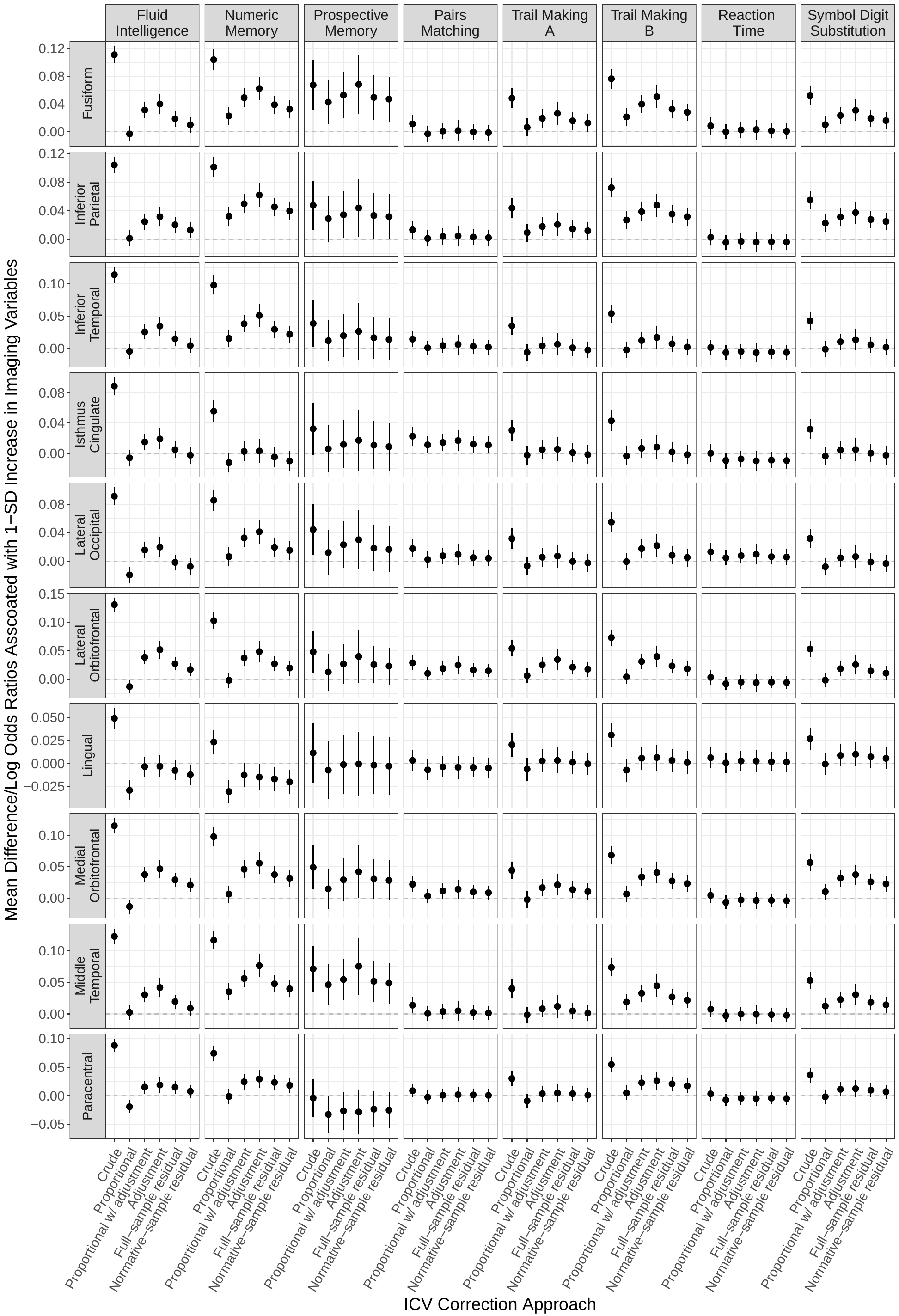

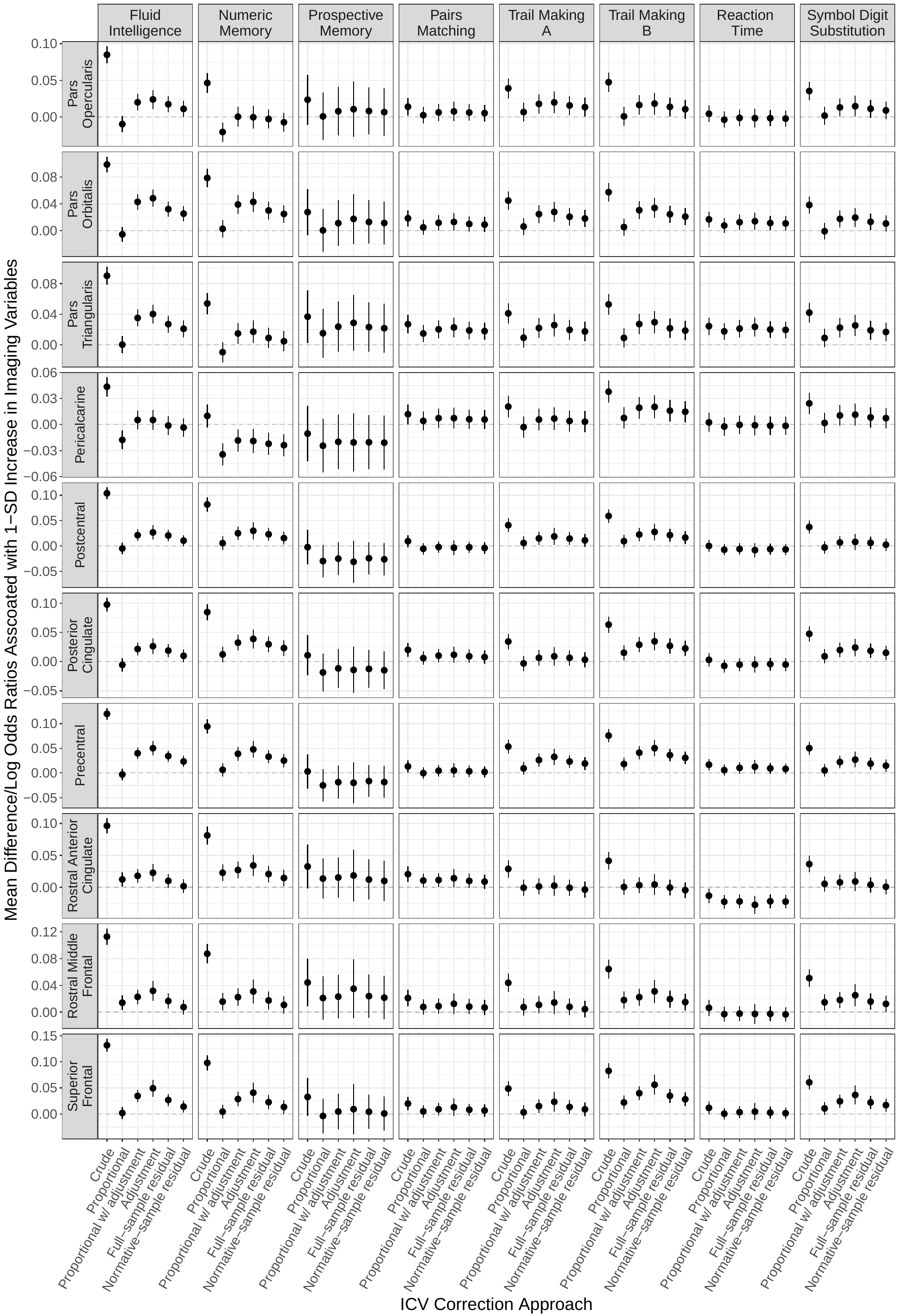

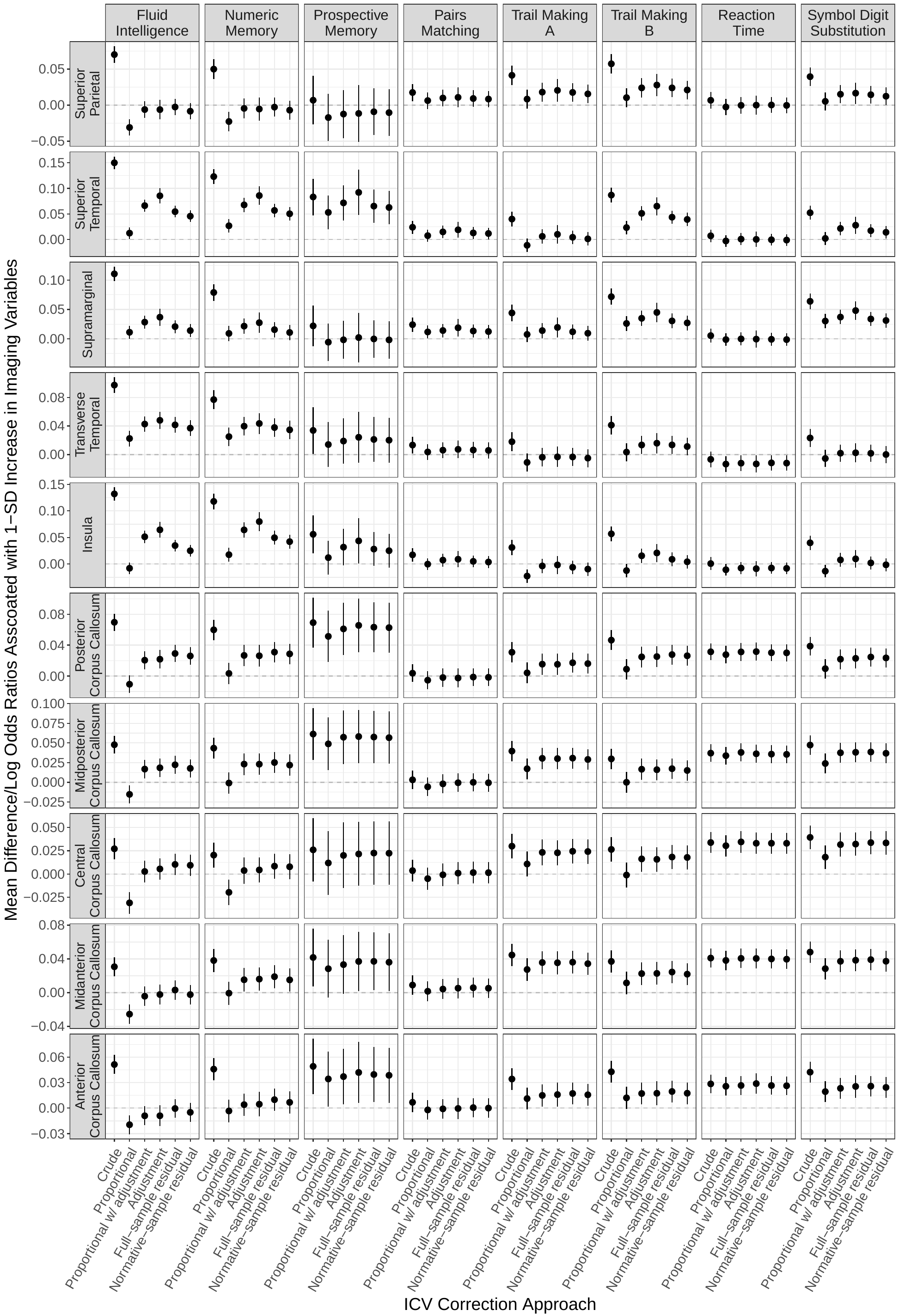

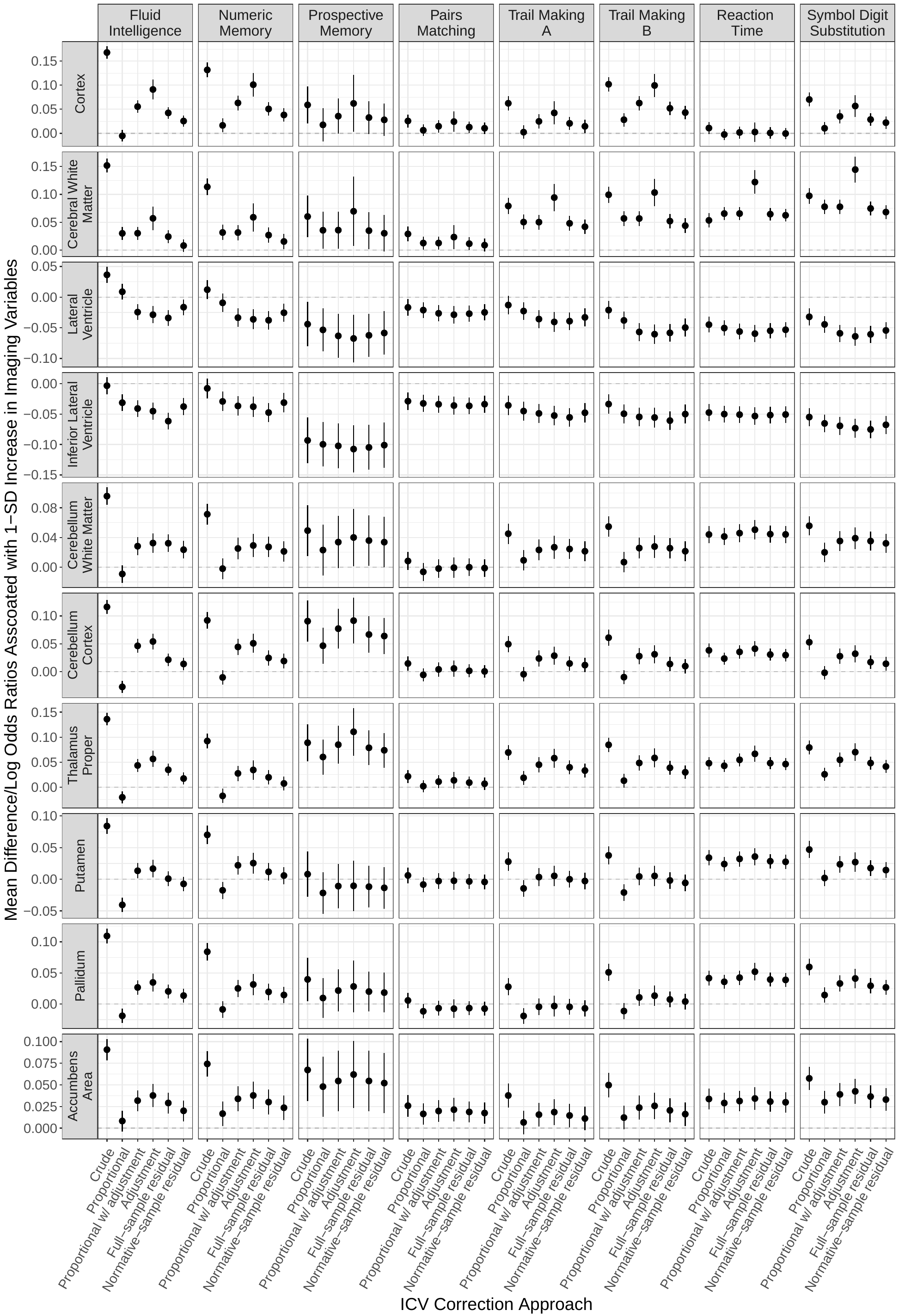

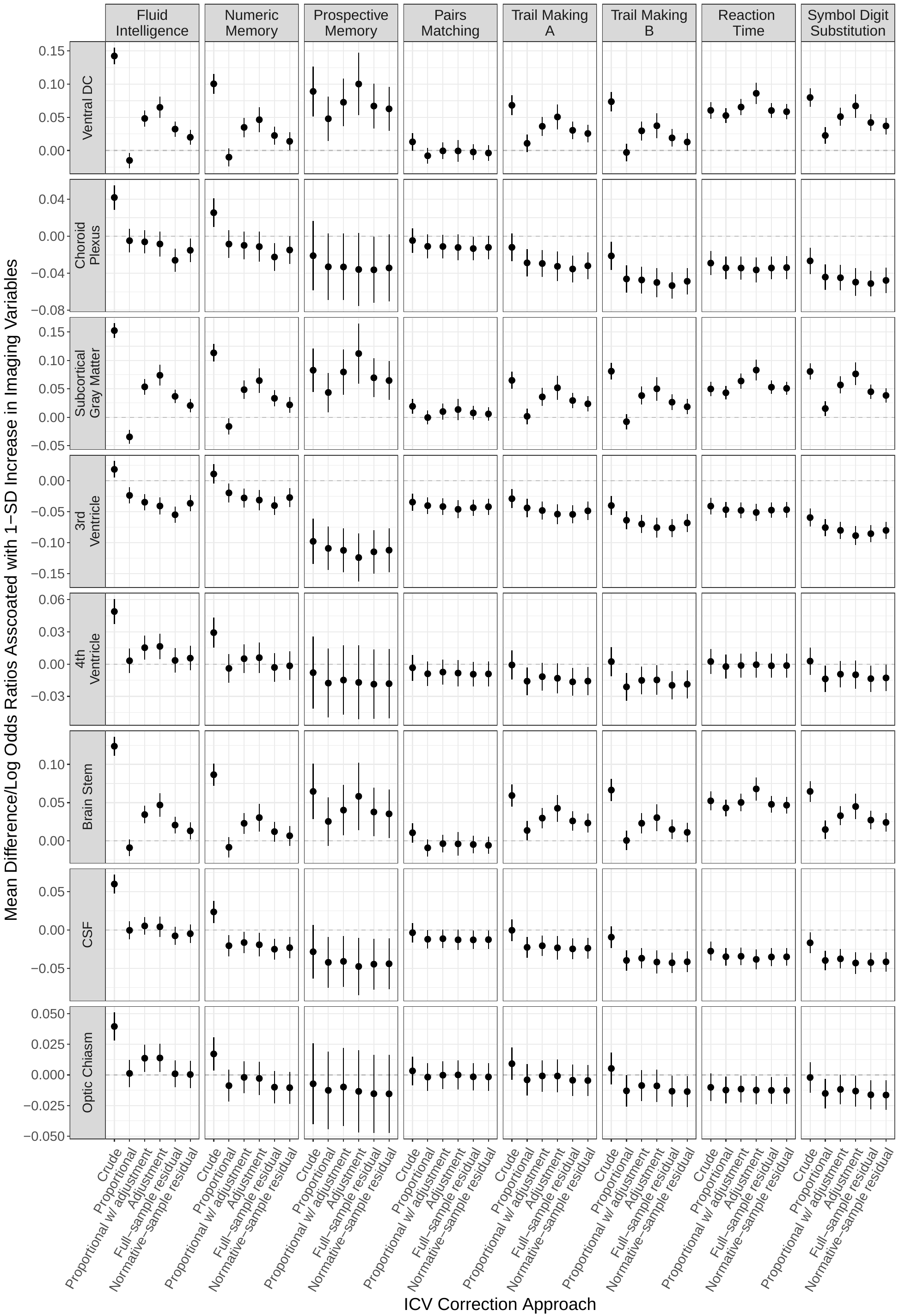

**Supplementary Figure S4: Pairwise correlations and consistencies for estimated associations between volumetric measures and A. fluid intelligence, B. numeric memory, C. prospective memory, D. pairs matching, E. Trails Making A, and F. Trails Making B, G. reaction time, and H. symbol digit substitution across all 58 brain regions assessed, excluding outliers of the regional volumes.**

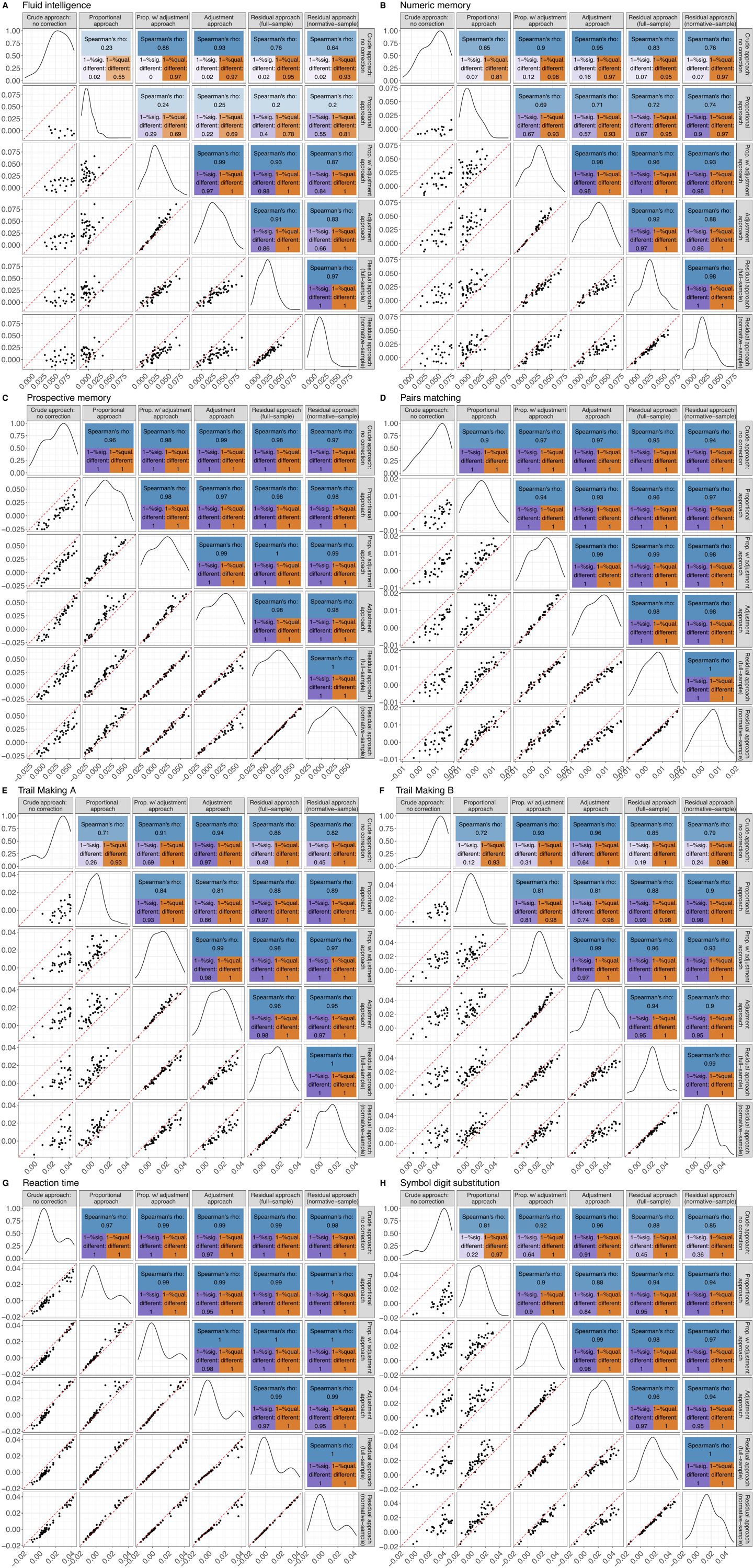

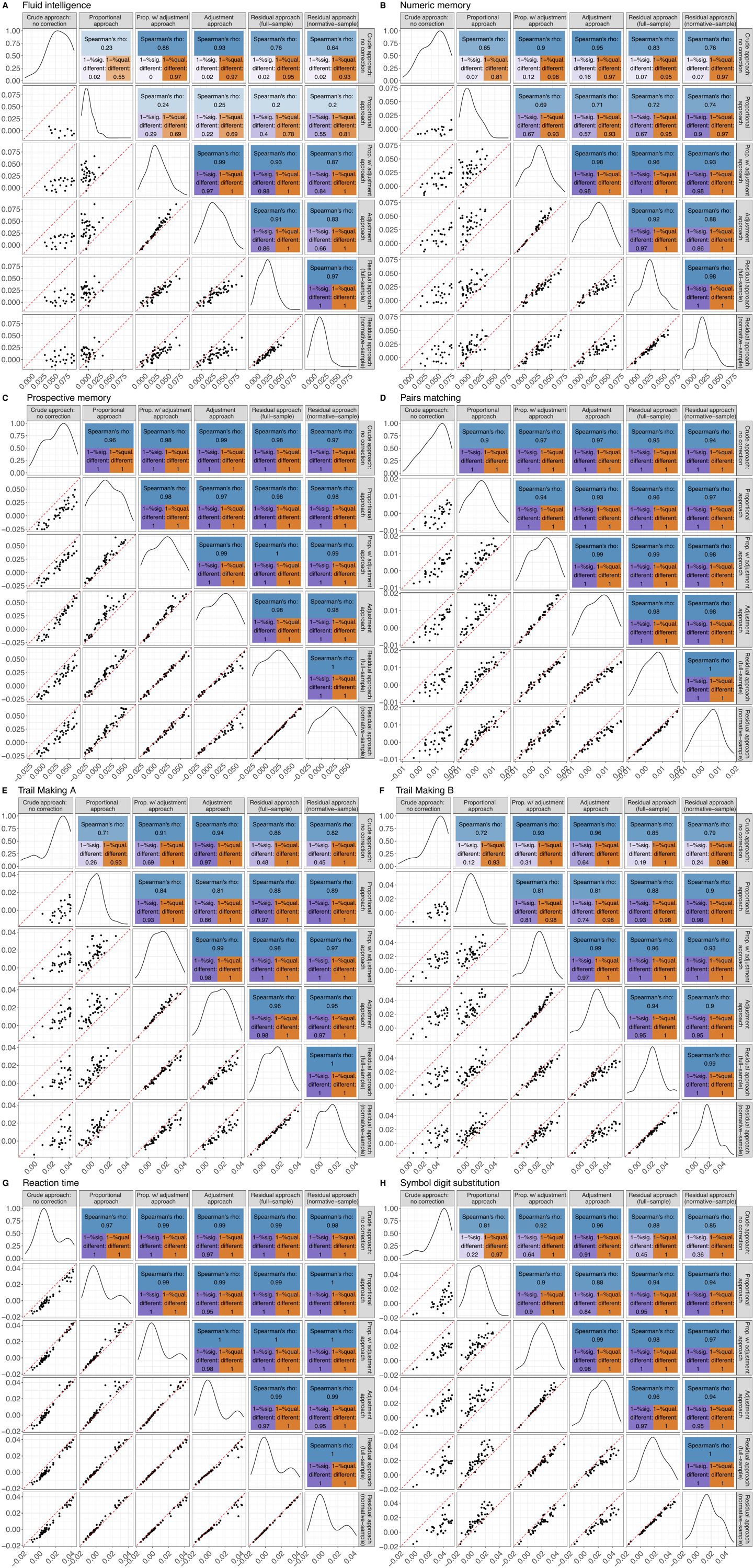

**Supplementary Figure S5: Associations between volumes of all regions and all cognitive scores, adjusting for age and age squared only.**
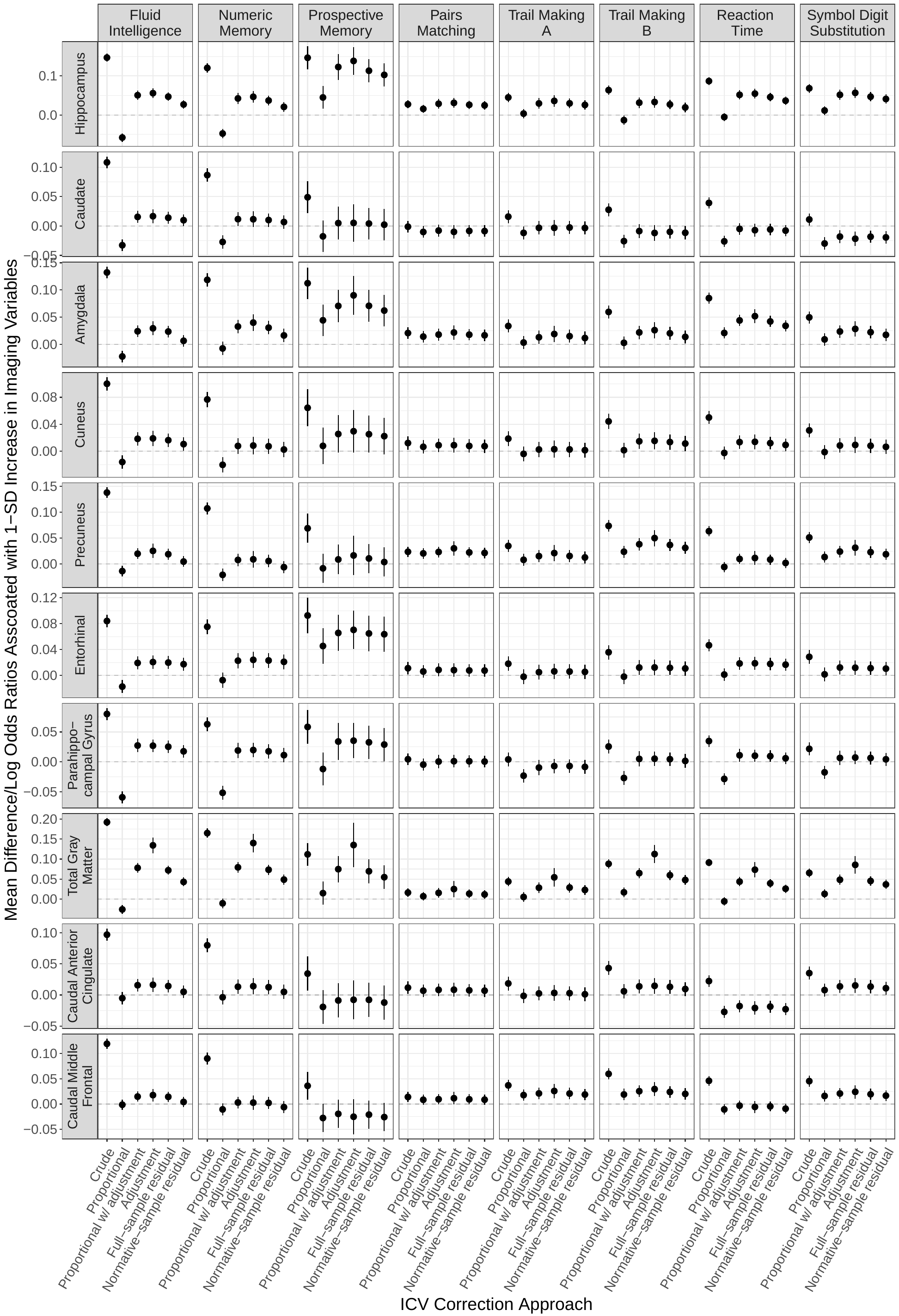

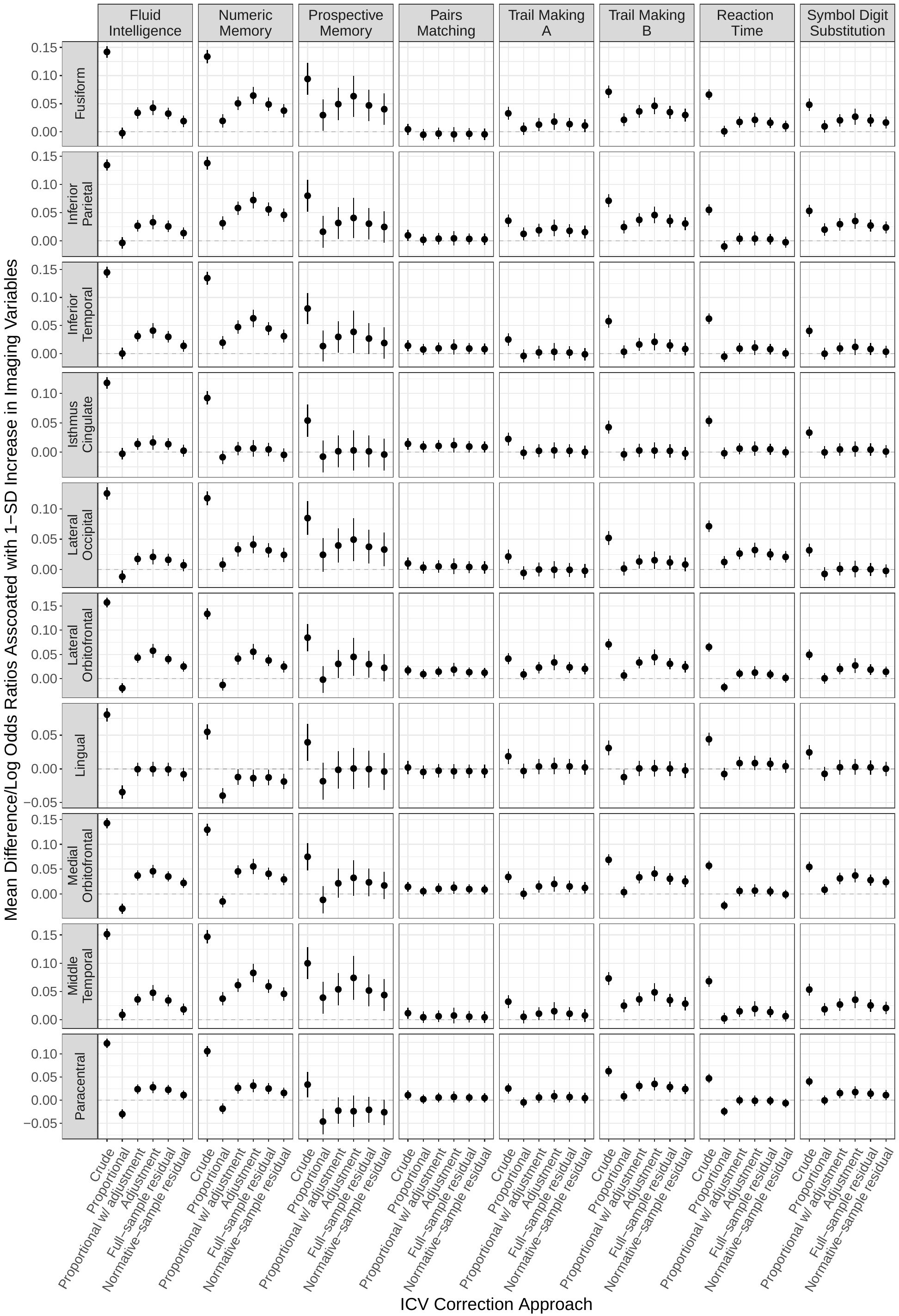

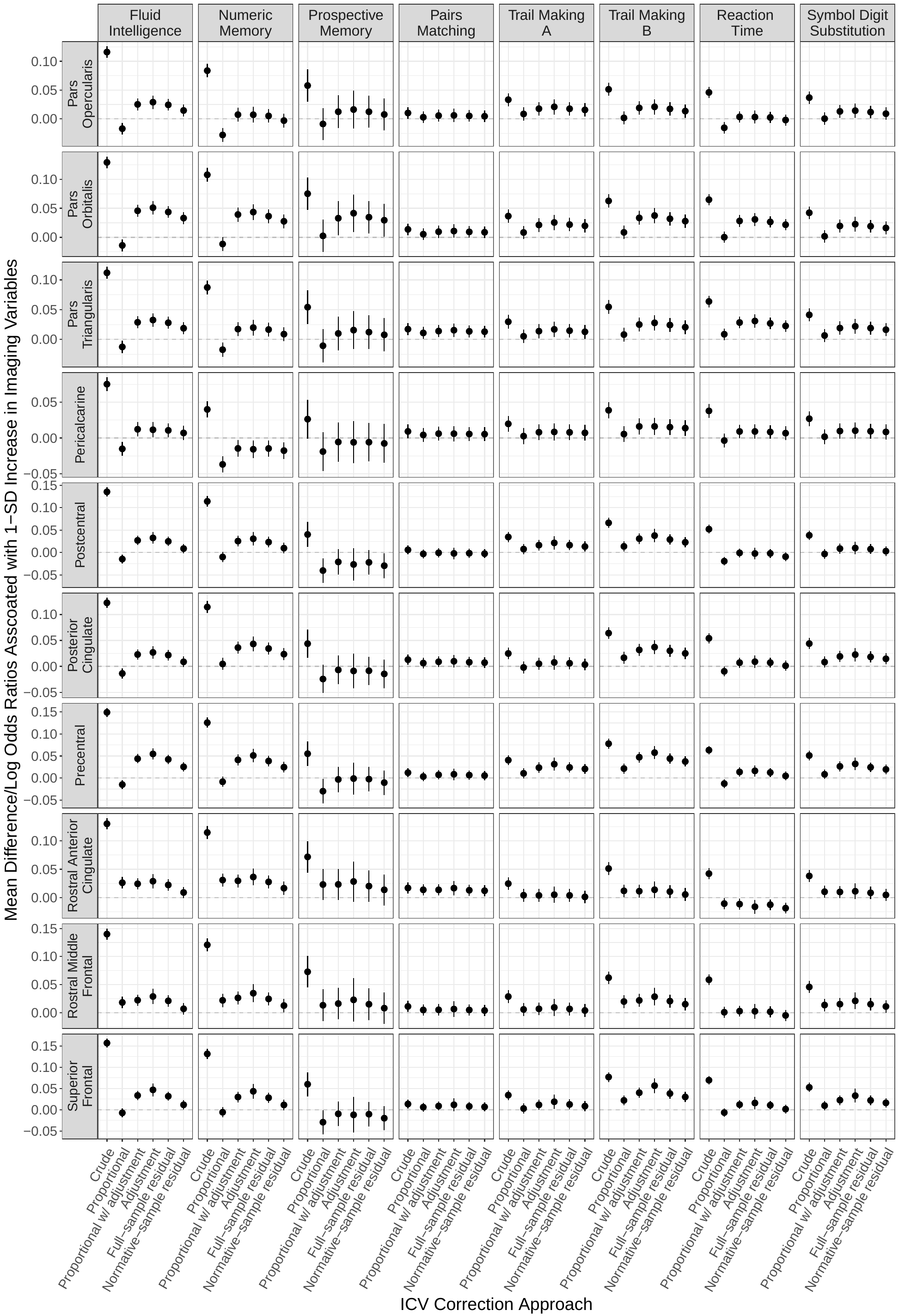

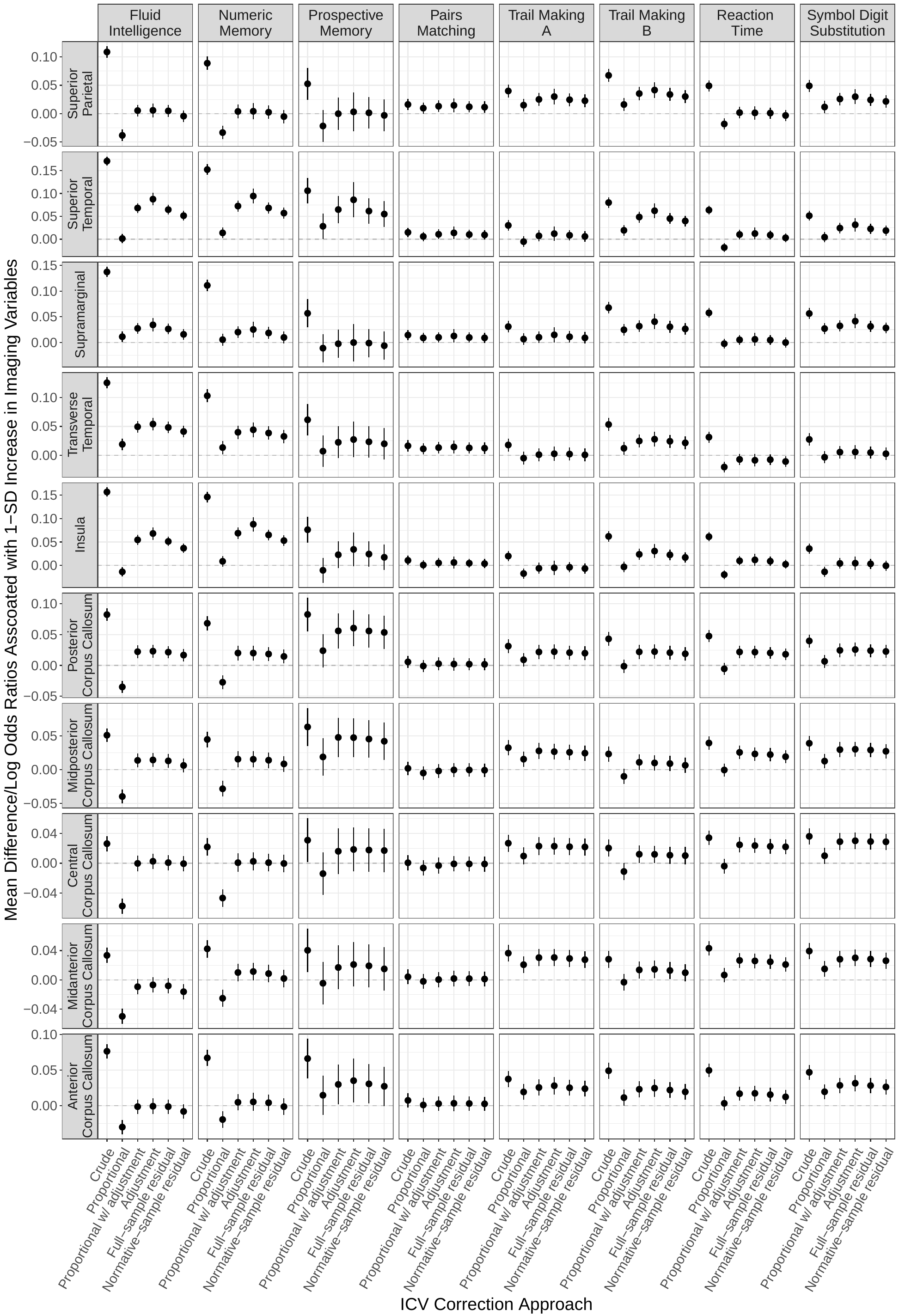

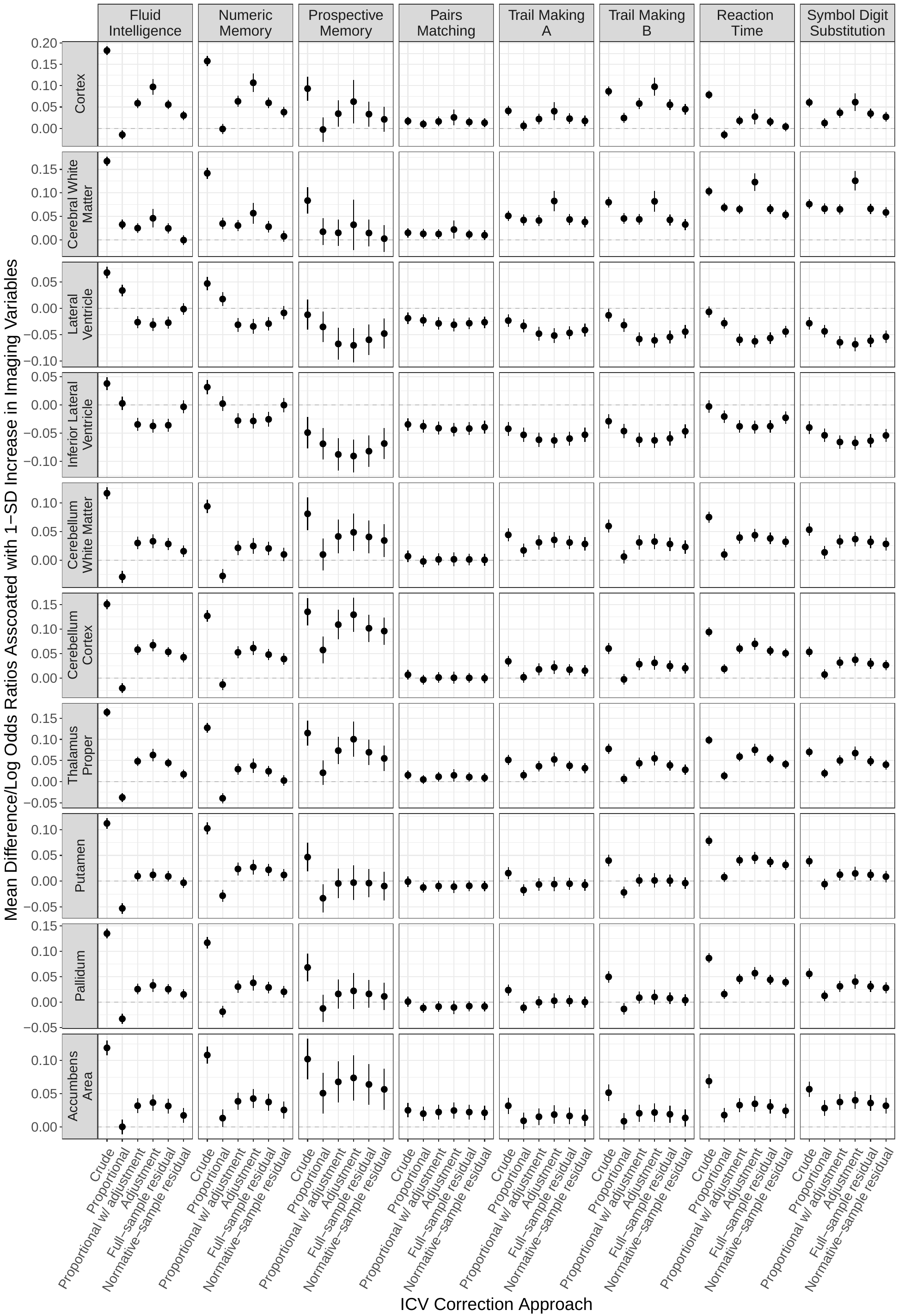

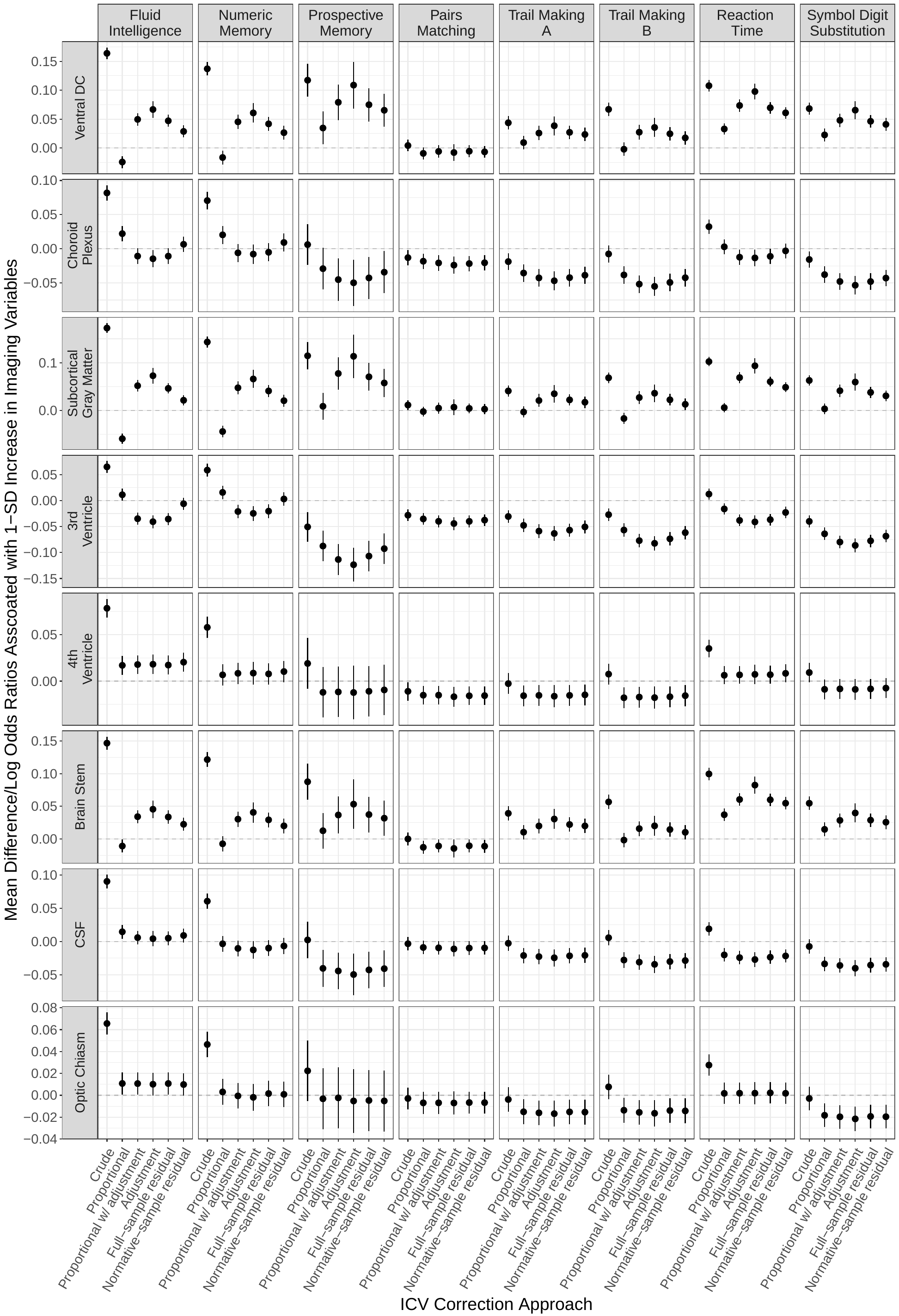

**Supplementary Figure S6: Pairwise correlations and consistencies for estimated associations between volumetric measures and A. fluid intelligence, B. numeric memory, C. prospective memory, D. pairs matching, E. Trails Making A, and F. Trails Making B, G. reaction time, and H. symbol digit substitution across all 58 brain regions assessed, adjusting for age and age squared only.**

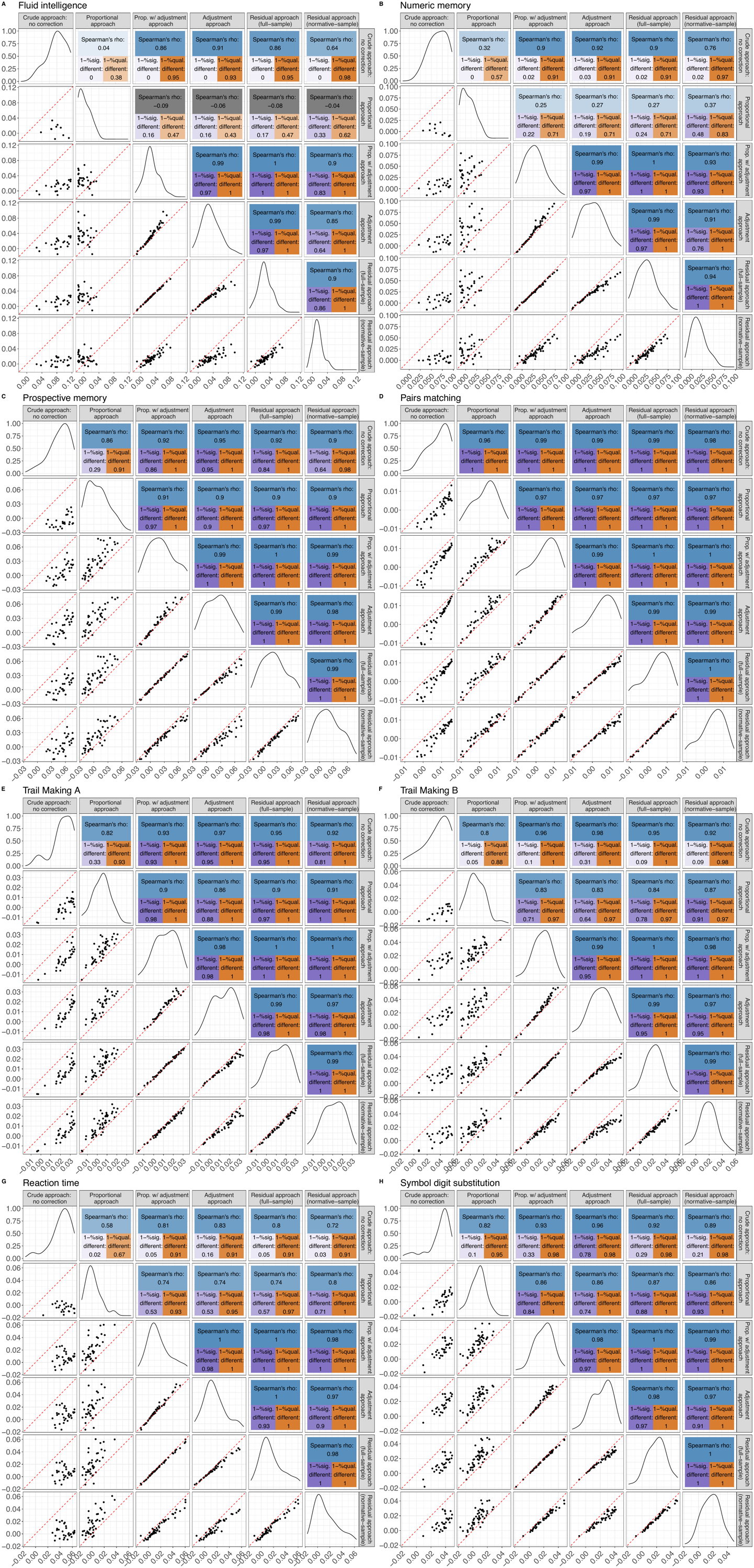

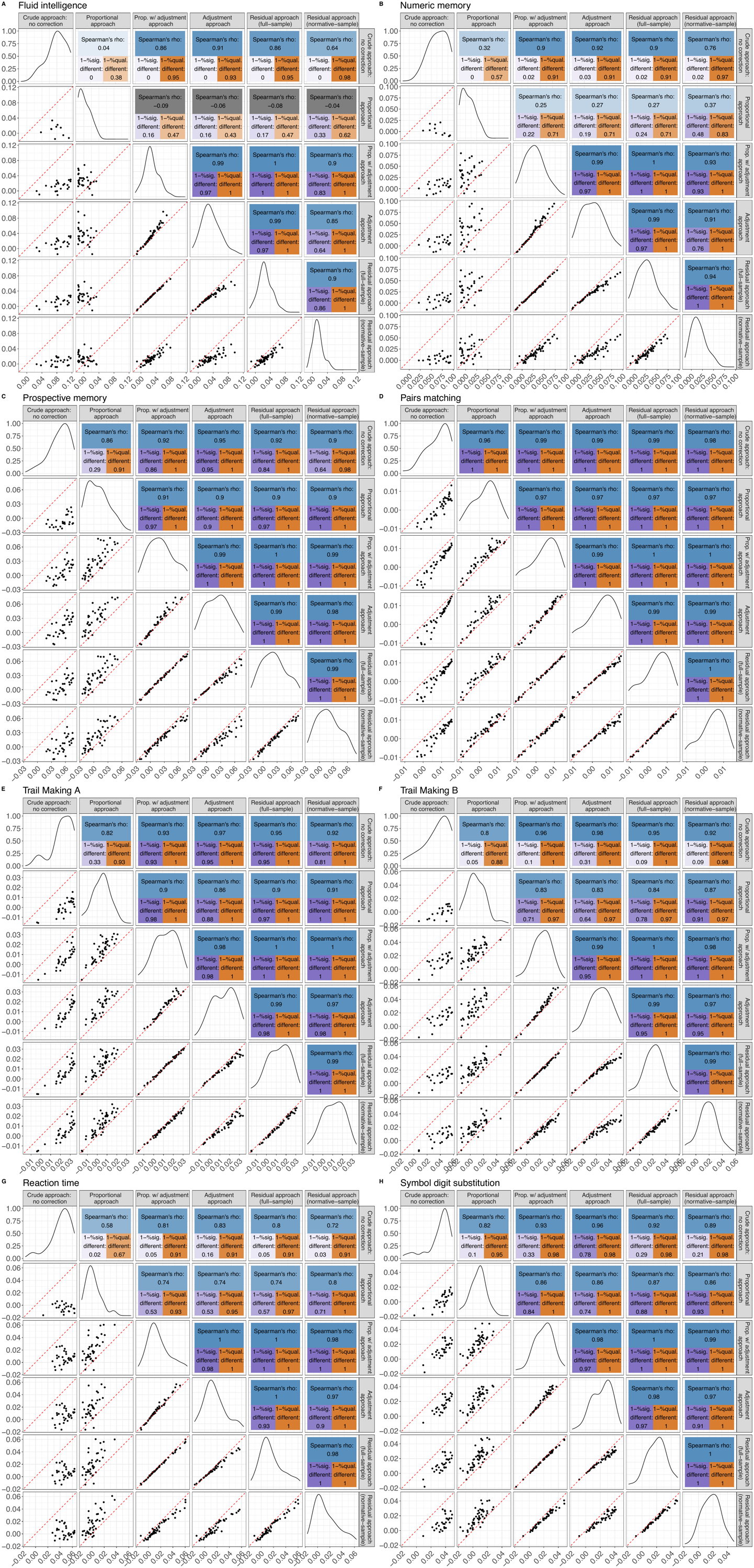

**Supplementary Figure S7: Associations between volumes of all regions and incident dementia, adjusting for age and sex.**

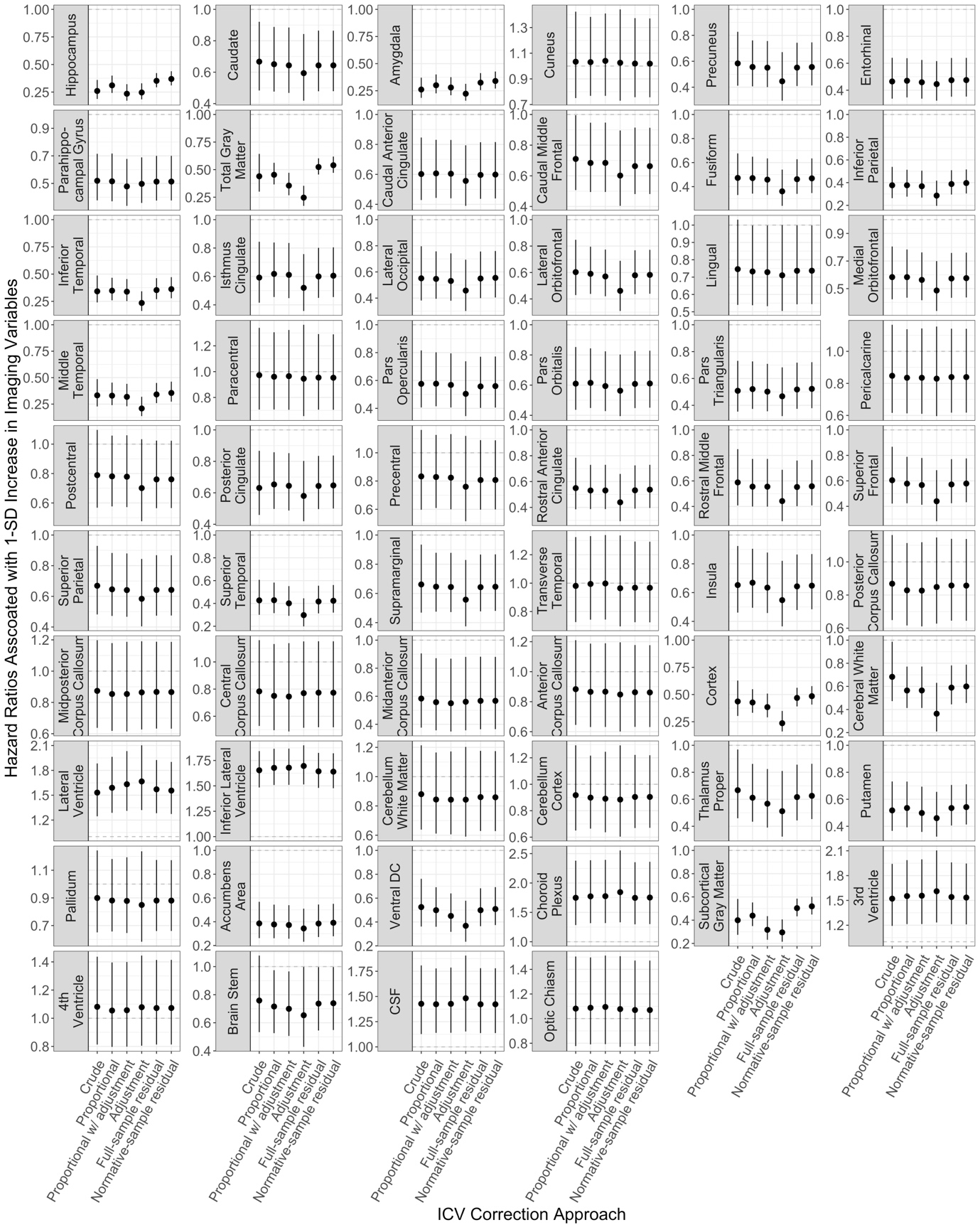
